# Capturing additional genetic risk from family history for improved polygenic risk prediction

**DOI:** 10.1101/2022.01.06.22268853

**Authors:** Tianyuan Lu, Vincenzo Forgetta, J Brent Richards, Celia MT Greenwood

**Affiliations:** Lady Davis Institute for Medical Research, Jewish General Hospital, Montreal, Canada; Quantitative Life Sciences Program, McGill University, Montreal, Canada; Department of Epidemiology, Biostatistics and Occupational Health, McGill University, Montreal, Canada; Department of Human Genetics, McGill University, Montreal, Canada; Department of Twin Research and Genetic Epidemiology, King’s College London, London, United Kingdom; Gerald Bronfman Department of Oncology, McGill University, Montreal, Canada

## Abstract

Family history of complex traits may reflect transmitted rare pathogenic variants, intra-familial shared exposures to environmental and lifestyle factors, as well as a common genetic predisposition. We developed a latent factor model to quantify trait heritability in excess of that captured by a common variant-based polygenic risk score, but inferable from family history. For 941 children in the Avon Longitudinal Study of Parents and Children cohort, a joint predictor combining a polygenic risk score for height and mid-parental height was able to explain ∼55% of the total variance in sex-adjusted adult height z-scores, close to the estimated heritability. Marginal yet consistent risk prediction improvements were also achieved among ∼400,000 European ancestry participants for 11 complex diseases in the UK Biobank. Our work showcases a paradigm for risk calculation, and supports incorporation of family history into polygenic risk score-based genetic risk prediction models.

## Introduction

Predicting phenotypic values of complex traits, or the risks and outcomes of complex diseases have strong implications in health care and biomedical research^1, 2^. In recent years, large-scale genome-wide association studies (GWASs) have characterized the genetic architecture of many complex traits and diseases^3^. Developing polygenic risk scores aggregating the effects of well-profiled genetic determinants has become possible^4, 5^. Polygenic risk scores have demonstrated the potential to improve risk stratification in large populations^6–9^, assist diagnosis and clinical differentiation^10–12^, and refine risk management and treatment strategies^13–15^.

However, most polygenic risk scores only capture linear additive effects of common genetic variants. Currently, most GWASs – even those based on the largest biobank studies – restrict consideration to genetic variants with a minor allele frequency *>* 0.1% or higher^3, 16^. Furthermore, extremely rare pathogenic variants with high penetrance are rarely detected with array-based genotyping and imputation^3, 17^. Further-more, powerful approaches to accurately model more complex non-linear effects (i.e. dominance effects) and interaction effects (i.e. gene-by-environment effects and gene-by-gene effects) in a high dimensional setting are scarce, particularly since these effects are in general weaker than the linear additive effects^3, 18^. Despite continuing methodological innovations in mining hidden heritability, these under-captured genetic effects likely prevent polygenic risk scores from achieving a further-improved predictive performance.

In contrast, family history information, such as parental measures of phenotypic values and disease records, provides an indirect measure of the overall genetic predisposition amongst relatives^19^. Although traditionally considered as a crucial risk factor for Mendelian diseases, family history has also shown added value in polygenic risk prediction^20–24^. For instance, individuals at an elevated level of risk for various types of cancer and cardiovascular diseases are more likely to be identified by assessments combining the family history of disease with polygenic risk scores^20–24^, compared to using polygenic risk scores or family history alone.

Nevertheless, creating accurate joint predictors may be challenging, since it requires modelling individual-level training data on phenotypes, genotypes, and family history information, as has been explored previously^7, 25–27^. This may not lead to effective prediction models if datasets containing all the required information are too small, particularly for diseases with a low prevalence in the population.

Therefore, in this work, we demonstrated both by theory and with examples, the improved predictions associated with a scheme for combining polygenic risk scores and parental disease histories. We approached this goal by inquiring what proportion of trait variance is captured by parental information and not by existing polygenic risk scores. Then, in comparison to using polygenic risk scores alone or predictors based only on parental trait measures or parental disease history, we evaluated the performance of these joint predictors in predicting adult height amongst 2,397 European ancestry children in the Avon Longitudinal Study of Parents and Children (ALSPAC) cohort^28, 29^, as well as in predicting risk for 11 complex diseases amongst ^∼^400,000 European ancestry participants in the UK Biobank^30^. An R toolkit implementing the method developed in this work, called FHPRS (Family History-assisted Polygenic Risk Score), is openly available at https://github.com/tianyuan-lu/PRS-FH-Prediction.

## Results

### Inference of under-captured genetic components in family history

We propose a conceptual latent factor model to account for genetic components that are not modelled by polygenic risk scores but could be inferred from parental history (**Methods**). Briefly, for a continuous polygenic trait (**Figure 1a**), we assume that its genetic determinants can be partitioned into two orthogonal genetic components: one component captured by a polygenic risk score used for prediction, and the other component representing under-captured genetic effects. The under-captured genetic component could include the effects of unmeasured common variants, rare variants, gene-by-environment interactions, epistasis, intra-familial shared environmental or lifestyle factors, etc. We suppose that these two genetic components are independently passed on from the parents to the children, and parental measures of the trait may partially inform the under-captured genetic component (**Methods** and **Figure 1a**). This model can be adapted for binary diseases, wherein the parental disease history may inform the underlying genetic liability (**Figure 1b**).

**Figure 1.**
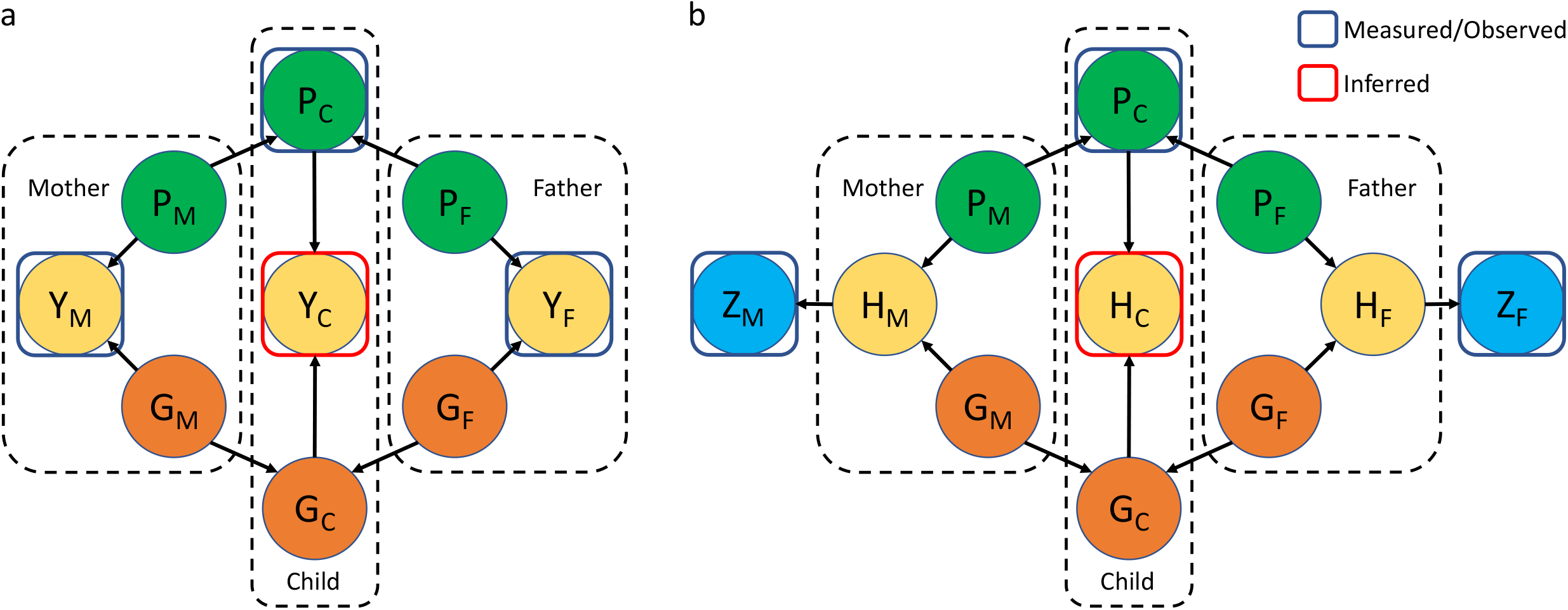
Causal diagrams representing latent factor models. (a) For the parents and the children separately, a continuous trait *Y* is determined by a polygenic component *P* captured by a polygenic risk score, and a latent genetic component *G* independent of *P*. (b) A binary trait *Z* is determined by the underlying genetic liability *H*. Analogous to *Y* in (a), *H* is jointly determined by *P* and *G*. Prediction combining a polygenic risk score and parental information is equivalent to inference of the distribution of *Y* in (a) and *H* in (b) for the children. The two genetic components, *P* and *G*, are assumed to be independently passed on from the parents to the offspring. Trait heritability is assumed to be constant across different generations.

Based on this model, a multivariate predictive model for a continuous trait or the genetic liability for a disease can be created by combining a polygenic risk score and family history, requiring only (1) the magnitude of association between a polygenic risk score and the target trait or disease, and (2) the magnitude of association between parents’ trait measures or disease history and the target trait or disease amongst children (**Methods**). Importantly, these estimates can be obtained from separate well-powered reference cohort studies, without the need to access individual-level information for training the predictive model. Subsequently, predictions for individuals in a test population or patients in clinics can be obtained based on their genotypes and parental trait measures or family disease history.

### Improved height prediction amongst children by incorporating parental height measures

We assessed the performance of this joint predictor in predicting children’s adult height. A polygenic risk score for age and sex-adjusted adult height z-score was recently developed using resources from the UK Biobank and the Genetic Investigation of ANthropometric Traits consortium^7, 30, 31^. On an out-of-sample test dataset in the UK Biobank, this score explained 36.7% of the total variance in height z-score^7^. To complement this score, an observational study found that the mid-parental height z-score was able to explain 44.9% of the total variance in offspring height z-scores in the Erasmus Rucphen Family Study^32^. Based on these estimates, we derived that 58.1% of the total height z-score variance was under-captured and could be partially inferred from parental height measures (**Methods**).

A joint predictor combining this polygenic risk score and mid-parental height was then obtained for European ancestry children in the ALSPAC cohort (**Methods** and **Supplementary Table 1**). Consequently, among 941 genotyped children who had both biological parents’ height measures, this joint predictor was able to explain 55.3% of the total variance in their sex-adjusted adult height z-score (**Figure 2a**). In contrast, similar to metrics obtained in literature, the polygenic risk score alone explained 38.2% of the total variance, while the mid-parental height z-score alone explained 43.1% (**Figure 2a**). As expected, the joint predictor also achieved the highest prediction accuracy with a root-mean-square error (RMSE) of 4.212 cm, compared to 4.955 cm by the polygenic risk score and 4.752 cm by the mid-parental height z-score (**Figure 2b**).

**Figure 2.**
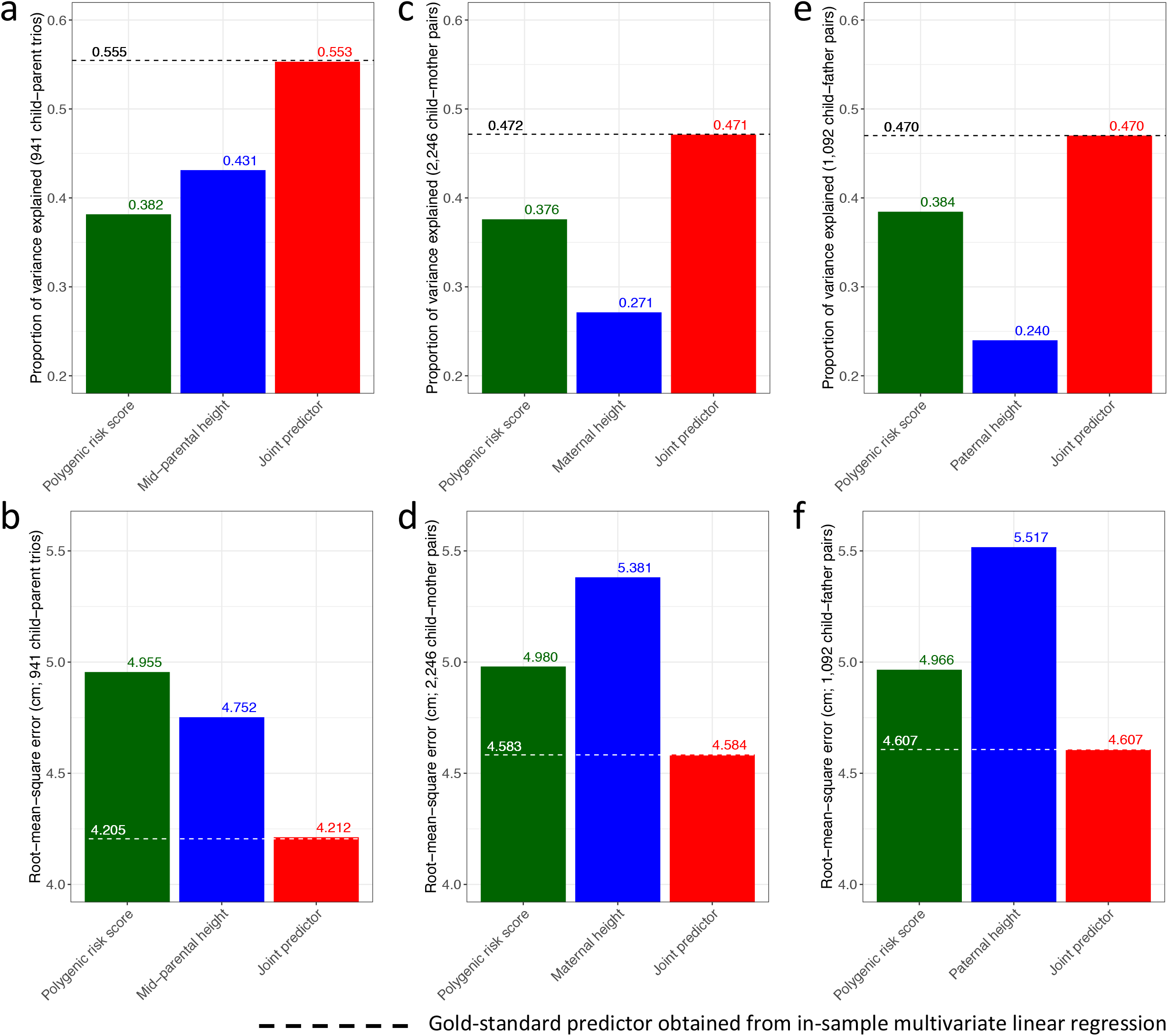
Comparison of predictive performance in predicting children’s sex-adjusted adult height z-scores for a polygenic risk score, parental height measures, and a joint predictor. A joint predictor combining the polygenic risk score and the mid-parental height predictor achieves (a) the highest proportion of variance explained, and (b) the lowest root-mean-square error based on 941 child-parent trios. A joint predictor incorporating (c) and (d) only the maternal height for 2,246 child-mother pairs, or (e) and (f) only the paternal height for 1,092 child-father pairs also outperforms the polygenic risk score and the single parental height predictor alone. These joint predictors have similar predictive performance as the corresponding gold-standard in-sample best linear unbiased predictors, indicated by dashed lines.

Furthermore, polygenic prediction could also be improved when only one parent’s height was measured (**Methods**). Specifically, among 2,246 children with maternal height measures, a joint predictor incorporating the polygenic risk score and maternal height z-score explained 47.1% of the total variance and achieved an RMSE of 4.584 cm (**Figure 2c and d**). Among 1,092 children with paternal height measures, a joint predictor incorporating the polygenic risk score and paternal height z-score explained 47.0% of the total variance and achieved an RMSE of 4.607 cm (**Figure 2e and f**). Evidently, these joint predictors demonstrated superior predictive performance over the polygenic risk score or parental height alone.

Notably, all joint predictors demonstrated almost identical predictive performance as an in-sample combination of the polygenic risk score and parental height z-scores obtained from multivariate linear regression using individual-level data (**Figure 2**).

### Improved complex disease risk prediction by incorporating parental disease history

Next, we tested whether a joint predictor could improve polygenic risk prediction for 11 complex diseases in the UK Biobank (**Supplementary Table 2**). Polygenic risk scores have been recently developed for each of these diseases and are documented in the PGS Catalog (**Supplementary Table 3**)^33^. Estimation of under-captured genetic effects was conducted based on a training dataset consisting of 10% of the participants from the UK Biobank (**Methods** and **Supplementary Figure 1**). For most of these complex diseases, individuals having a parental disease history were significantly more likely to have the corresponding disease (**Supplementary Table 4**). The magnitudes of association reflect that a substantial proportion of genetic influence is not captured by the polygenic risk scores (**Supplementary Table 4**).

Incorporation of parental disease history into polygenic risk predictions led to a re-stratification of the predicted risks (**Figure 3a**). Specifically, individuals with a parental disease history would be considered at an elevated level of risk compared to those with a similar polygenic risk score but without a parental disease history. The discriminative power of polygenic risk scores in identifying individuals who developed the corresponding diseases was significantly improved (**Table 1** and **Supplementary Table 5**). For instance, combined with age, sex, genotyping array, recruitment centre, and the first 10 genetic principal components, a joint predictor for myocardial infarction achieved an area under the receiver operating characteristic curve (AUROC) of 0.7625 (**Figure 3b**) and an area under the precision-recall curve (AUPRC) of 0.0834 (**Figure 3c**), based on a test dataset consisting of 90% of the participants from the UK Biobank (**Methods** and **Supplementary Figure 1**). In contrast, prediction based on the polygenic risk score (with other covariates but without parental history) had an AUROC of 0.7567 (DeLong’s test p-value = 5.3 × 10^−13^) and an AUPRC of 0.0800; prediction based on the parental disease history (with other covariates but without the polygenic risk score) had an AUROC of 0.7375 (DeLong’s test p-value = 3.0 × 10^−126^) and an AUPRC of 0.0671 (**Figure 3b** and **c**).

**Figure 3.**
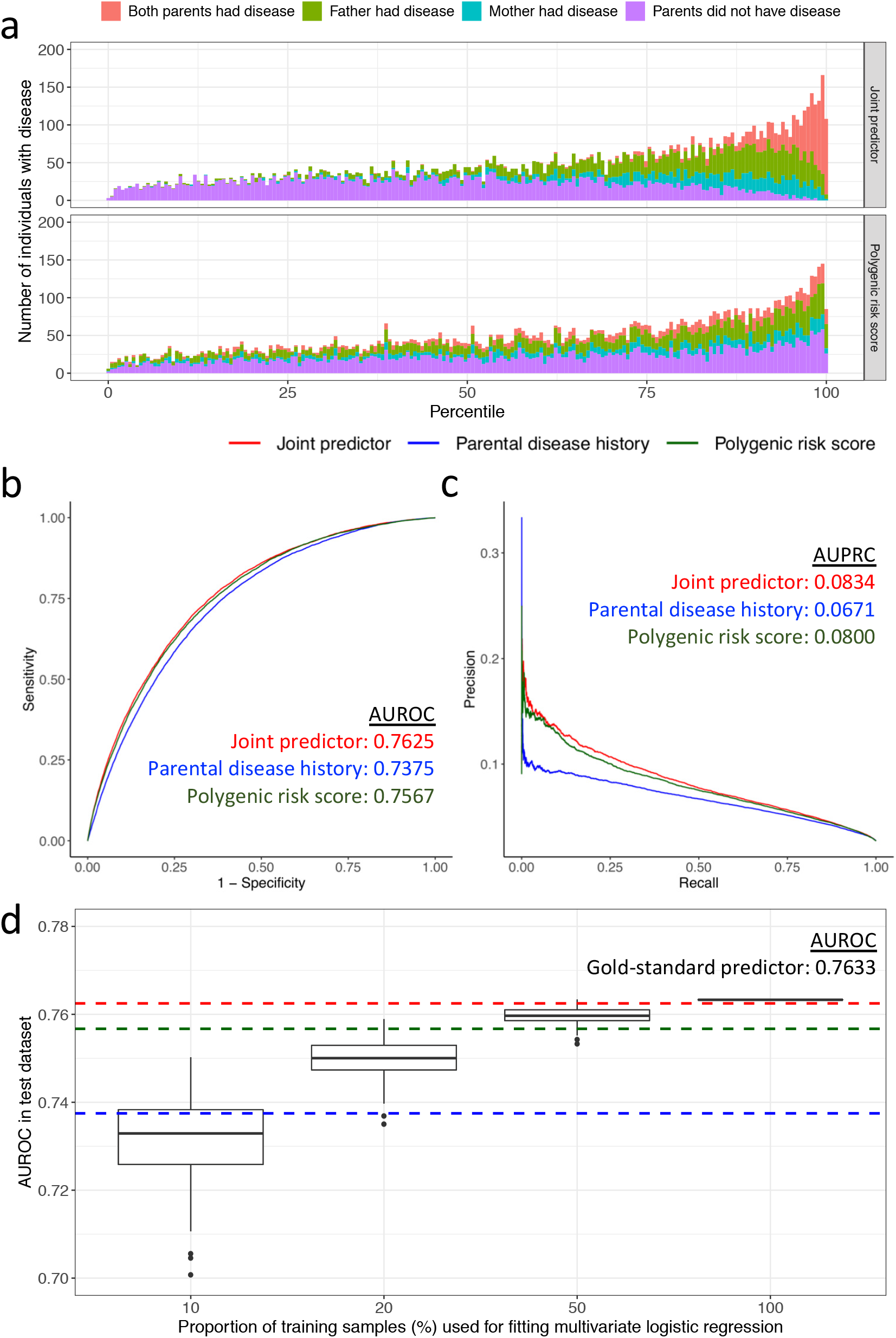
A joint predictor improves risk stratification for myocardial infarction in the UK Biobank. (a) Incorporation of parental disease history leads to calibration of polygenic risk scores. Individuals with a parental disease history are more likely to be considered at risk. The joint predictor achieves higher (b) area under the receiver operating characteristic curve (AUROC) and (c) are under the precision-recall curve (AUPRC), compared to the polygenic risk score and the parental disease history. (d) Performance of a data-driven joint predictor obtained from multivariate logistic regression is sensitive to sample size when using individual-level data. Box plots represent distributions of AUROC obtained in 100 replicates corresponding to different sample sizes. AUROC of the joint predictor, polygenic risk score, and parental disease history are indicated by red, green, and blue dashed lines, respectively.

**Table 1.** Improved discriminative power of a joint predictor in identifying individuals at an elevated risk of disease based on the UK Biobank test dataset.

| Disease | AUROC<sup>a</sup> (p-value of DeLong's test<sup>b</sup>) |  |  | NRI<sup>c</sup> (%) at score percentile cutoff |  |  |  | IDI<sup>c</sup> (%) |
| --- | --- | --- | --- | --- | --- | --- | --- | --- |
|  | Polygenic risk score | Parental history | Joint predictor | 50 | 80 | 95 | 99 |  |
| Breast cancer | 0.6443<br>(5.8x10<sup>-16</sup>) | 0.5771<br>(8.9x10<sup>-213</sup>) | 0.6500 | +0.94 | +0.84 | +0.14 | +0.17 | +0.56 |
| Prostate cancer | 0.7567<br>(1.4x10<sup>-3</sup>) | 0.7071<br>(2.6x10<sup>-197</sup>) | 0.7588 | +0.29 | +0.24 | +0.68 | +0.12 | +0.21 |
| Colorectal cancer | 0.6755<br>(5.4x10<sup>-8</sup>) | 0.6676<br>(2.1x10<sup>-41</sup>) | 0.6768 | +0.27 | +0.64 | +0.45 | 0 | +0.13 |
| Lung cancer | 0.6915<br>(1.6x10<sup>-7</sup>) | 0.6947<br>(1.3x10<sup>-6</sup>) | 0.6975 | +2.46 | +5.07 | +3.01 | +1.12 | +2.60 |
| Myocardial infarction | 0.7567<br>(5.3x10<sup>-13</sup>) | 0.7375<br>(3.0x10<sup>-126</sup>) | 0.7625 | +0.85 | +0.70 | +0.96 | +0.09 | +0.58 |
| Ischemic heart disease | 0.7445<br>(1.5x10<sup>-38</sup>) | 0.7398<br>(3.4x10<sup>-297</sup>) | 0.7520 | +1.10 | +0.96 | +0.57 | +0.22 | +0.75 |
| Stroke | 0.6862<br>(6.2x10<sup>-10</sup>) | 0.6802<br>(8.7x10<sup>-16</sup>) | 0.6870 | +0.41 | +0.06 | +0.38 | +0.14 | +0.49 |
| Type 2 diabetes | 0.7129<br>(6.7x10<sup>-31</sup>) | 0.7072 (< 1.0x10<sup>-350</sup>) | 0.7266 | +1.71 | +2.50 | +1.47 | +0.89 | +1.37 |
| Alzheimer's disease | 0.7571<br>(1.4x10<sup>-7</sup>) | 0.7616<br>(1.5x10<sup>-3</sup>) | 0.7645 | +4.48 | +6.65 | +5.76 | +2.18 | +4.02 |
| Parkinson's disease | 0.7482<br>(2.6x10<sup>-2</sup>) | 0.7412<br>(2.1x10<sup>-13</sup>) | 0.7507 | +1.86 | +2.21 | +0.59 | +1.08 | +1.22 |
| COPD | 0.7325<br>(4.4x10<sup>-58</sup>) | 0.7448<br>(2.0x10<sup>-11</sup>) | 0.7463 | +7.03 | +11.88 | +5.07 | +1.60 | +5.96 |

<sup>a</sup> Including effects of age, sex (except for breast cancer and prostate cancer), genotyping array, recruitment centre, and the first 10 genetic principal components of ancestry
<sup>b</sup> Comparing AUROC of the polygenic risk score or parental history to that of the joint predictor
<sup>c</sup> Comparing the joint predictors to the corresponding polygenic risk scores; a positive number indicates improved re-classification

Meanwhile, the joint predictor could more accurately assign individuals into higher-vs-lower risk groups, indicated by positive net-reclassification indices (NRI) across different percentile cutoffs and positive integrated discrimination improvement (IDI) indices (**Table 1**). For example, at the 95-th percentile cutoff (i.e. 5% of the population to be considered at high risk), the joint predictor for Alzheimer’s disease had the highest NRI of 5.76% over the polygenic risk score for Alzheimer’s disease, followed by the joint predictor for chronic obstructive pulmonary disease (COPD), with an NRI of 5.07% (**Table 1**). Notably, the joint predictor for Alzheimer’s disease was significantly associated with carrying one or two *APOE* e4 alleles (**Methods** and **Supplementary Figure 2a** and **b**), which is a well-known genetic risk factor for Alzheimer’s disease^34–36^ but is not included in the polygenic risk score for this analysis. In addition, individuals with a higher score in the joint predictor for COPD were slightly yet significantly more likely to be ever-smokers, while the the polygenic risk score alone did not capture the genetic predisposition to smoking (**Methods** and **Supplementary Figure 2c**).

For most diseases under investigation, the joint predictors consistently achieved comparable predictive performance to gold-standard predictors obtained by fitting multivariate logistic regression models based on individual-level data from the training datasets (**Methods** and **Supplementary Figure 1**). For breast cancer, lung cancer, stroke and Alzheimer’s disease, the joint predictors even achieved a marginally higher AUROC than the data-driven predictors (**Table 1** and **Supplementary Figure 3**), though the joint predictors for ischemic heart disease and type 2 diabetes did not appear to be ideal (**Table 1** and **Supplementary Figure 3**). However, as expected, all of the individual-level data-driven predictors demonstrated sensitivity to sample size (**Figure 3d** and **Supplementary Figure 3**). For instance, for myocardial infarction, if a regression-based predictor were to be derived using ≤ 20% of the individuals included in the current training dataset, its discriminative power would almost certainly be worse than the polygenic risk score alone (**Figure 3d**).

## Discussion

Polygenic risk scores for complex traits are effective research tools and may greatly improve personalized health care in clinical practice^1, 2^. Development of polygenic risk scores relies on accurate characterization of genetic determinants in large-scale GWASs. Due to limitation of resources, most existing polygenic risk scores are restricted to modelling linear additive effects of common genetic variants, thus may benefit from being combined with predictors that are able to capture more elusive genetic effects. In this study, we have proposed a simple latent factor model to quantify and extract heritability not captured by polygenic risk scores but inferable from family history. We have systematically investigated the utility of adding family history into polygenic risk score-based polygenic risk prediction models.

The combination of parental height measures and a polygenic risk score for height brought substantial improvements in adult height prediction. In fact, the proportion of variance explained in adult height z-scores by our joint predictor in the ALSPAC cohort was similar to the estimated total SNP heritability of height z-scores^37–40^. On the other hand, for 11 complex diseases, the joint predictors consistently demonstrated stronger discriminative power in identifying individuals at an elevated level of risk, although the improvements appeared to be marginal. This may reflect limited sensitivity of relevant metrics to addition of new predictors^41^, as we observed in simulation studies (**Supplementary Note 1** and **Supplementary Figure 4**). Nonetheless, by jointly examining all metrics (AUROC, AUPRC, NRI, and IDI) as well as the distribution of predicted risks, we posit that the resulting risk re-stratification could still benefit up to thousands of individuals at the biobank-scale.

In developing this prediction scheme, we partitioned trait or disease heritability onto two orthogonal genetic components, where family history is assumed to partially inform a latent genetic component despite being correlated with the polygenic risk score. To account for the polygenicity and complexity of the underlying genetic effects, the genetic components were assumed to be normally distributed. We recognize that while these assumptions have strength in facilitating model specification and construction of joint predictors, they may not be theoretically optimal. For example, if strong gene-by-environment interaction effects exist, the under-captured genetic component should, at least in part, be correlated with the polygenic risk score. However, correlation or interaction between the two genetic components is unidentifiable without accessing individual-level training data. Encouragingly, our simulation studies indicated that our method could tolerate mild-to-moderate violations of model assumptions as well as errors in estimating model parameters (**Supplementary Notes 2** and **3**, and **Supplementary Figures 5-10**). Moreover, despite these strong assumptions, we found in practice that polygenic prediction could consistently be improved through our models built with this point of view, regardless of the underlying genetic architecture of complex traits, disease prevalence, and the methods adopted to develop polygenic risk scores^33^. In contrast to multivariate regression-based joint predictors, our method can leverage results of association tests obtained from separate cohort studies with a high statistical power, thus this method does not suffer when large individual-level training datasets are unavailable.

Our findings have important implications for developing polygenic risk predictors, since the undercaptured genetic components may have known sources. For instance, a large proportion of Alzheimer’s disease risk heritability is conferred by the common allele of *APOE* e4^34–36^. Not surprisingly, the parental disease history of Alzheimer’s disease appeared to be more predictive of the disease risk compared to a polygenic risk score not including this allele, while the joint predictor demonstrated clear advantages in risk stratification. Furthermore, by design, inherited risk factor exposure was also included in the under-captured genetic component. Smoking is one of the most important risk factors for COPD^42^ and is heritable^43, 44^. In our model, family disease history of COPD may partially capture the genetic predisposition to smoking as well as other risk factors that is not fully represented in the polygenic risk score. As a result, the corresponding joint predictor aggregating additional genetic risks demonstrated a prominent improvement over the polygenic risk score. These results not only support the utility of family history in enhancing predictive power, but also encourage explicit modelling of large effects such as monogenic causes or significant intra-familial shared risk factor exposures in polygenic risk prediction.

We note possible model mis-specification for ischemic heart disease and type 2 diabetes because the joint predictors we constructed displayed compromised predictive performance compared to the gold-standard predictors. We hypothesize that this may be due to errors in the definitions of disease in the parental histories as well as in the medical histories of the participants. Specifically, sub-classifications of diseases were not available for parental heart disease and diabetes, where the former may include various types of diseases affecting the cardiovascular system, and the latter may include both type 1 and type 2 diabetes. Consistent disease definitions and comprehensive phenotyping are thus required for validating our findings in clinical practice and in research.

We anticipate that the predictive performance of the joint predictors could be further enhanced with additional knowledge of family history, such as the disease history of relatives other than the parents, using empirical genetic relatedness based on pedigree information. However, we expect that information gained from second-degree or more distant relatives would be less significant compared to first-degree relatives. Furthermore, in the UK Biobank, the parental disease history largely reflected the parents’ lifetime risk given that the participants (children) were aged above 40 years upon recruitment. Hence, appropriate modelling of age-dependent risks should be pursued for most complex diseases that do not have an early onset, if disease history of younger relatives were to be considered.

Last, our findings should be considered specific to the study populations. Participants in our study cohorts are predominantly of European ancestries, yet it has been widely recognized that polygenic risk scores can have largely attenuated predictive performance when applied to populations of different genetic ancestries^45, 46^. Hence, we recommend extensive validations of our results in diverse populations as well as development of ancestry-specific polygenic risk scores. Adding to this restriction, we also note that participants in the UK Biobank have been shown to be slightly healthier, less obese, and less likely to smoke and drink alcohol than the general population in the United Kingdom^47^. In particular, the average age of the study cohort was substantially younger than the average age of onset for Alzheimer’s disease and Parkinson’s disease^48, 49^, thus the corresponding disease prevalence was low. Therefore, if a joint predictor were constructed for other populations, we strongly recommend estimating model parameters based on reference cohort studies with similar demographic characteristics and prevalence of disease. This may be particularly important for large cohort studies and population-level screening programs where family history information is less comprehensive than in clinical settings.

In summary, we have developed a risk calculation scheme by incorporating family history into polygenic risk scores. We found a substantial proportion of complex trait heritability was under-captured and could be partly inferred from family history. Our findings support the utility of combining family history with polygenic risk scores as well as investigations into complex genetic effects not captured by existing polygenic risk scores for improved genetic risk prediction.

## Methods

### Related methods

Combining a genetic risk score with measures of traits or disease status of relatives is not unprecedented. The most straightforward approach is to fit a multivariate regression model including both the genetic risk score and family history variables as predictors^7, 25^. Alternatively, extensions of the best linear unbiased prediction (BLUP) method have been proposed by appending an empirical relatedness matrix based on pedigree information to a genotype-based relatedness matrix for modelling random effects in a mixed model setting^26, 27^. Using a similar framework, family history may also improve the power of genetic association tests^19^. However, these methods require access to a training dataset that simultaneously contains phenotypes, genotypes, and family history information. For modelling disease outcomes, a large number of cases is needed to ensure statistical power. This is often unlikely due to confidentiality restrictions or logistical constraints. In addition, the variance-covariance structure of genetic components specified in our method is also similar to that implemented in a few methods using family disease status to modify genetic risk estimates in known risk loci^50–52^. Nonetheless, these methods require a pre-specified estimate of disease heritability, which may be prone to error if no reference for the targeted population is available or if unmeasured covariate effects are not properly accounted for.

### A latent factor model for polygenic inheritance in parent-child trios

We first consider a simple latent factor model for a normally distributed polygenic trait

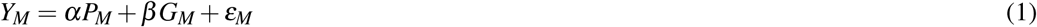

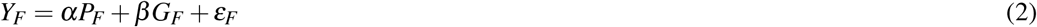

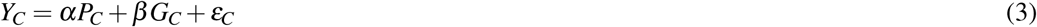

where the subscripts *M, F*, and *C* stand for the populations of mothers, fathers, and children, respectively. The mothers and the fathers are assumed to be unrelated. *P* represents the genetic component already captured by a polygenic risk score, while *G* represents the under-captured genetic component, including under-captured linear additive effects, non-additive effects, rare pathogenic variant effects, gene-gene or gene-environment interaction effects, inherited exposure to risk factors, etc. Importantly, *G* is assumed to be independent of *P*, and these two components are independently passed on from the parents to the children. We realize that this may be an unrealistic assumption, particularly for common genetic effects that were not captured in existing polygenic risk scores, but this assumption enables conceptual development of our latent model. Results in **Figure 2** and **Table 1** justify the usefulness of the perspective. *ε* captures the non-genetic residual variance in *Y*, where *ε* ⊥ (*P, G*). We assume both *P* and *G* are normally distributed because *P*, under a polygenic model, consists of multiple independent effects, each having an infinitesimal effect size, while *G* has a multifactorial nature. Without loss of generality, we suppose that *Y, P*, and *G* are scaled to have zero mean and unit variance.

We further assume that the effects of the modelled polygenic component and the under-captured genetic component, *α* and *β*, are invariant in the populations of parents and children from the same study. Naturally, the overall heritability of this trait is *α*^2^ + *β*^2^.

Next, we describe the joint distribution of ***P, G***, and ***Y***, where ***P*** = (*P*_*M*_, *P*_*F*_, *P*_*C*_)^⊤^, ***G*** = (*G*_*M*_, *G*_*F*_, *G*_*C*_)^⊤^, and ***Y*** = (*Y*_*M*_,*Y*_*F*_, *Y*_*C*_)^⊤^:

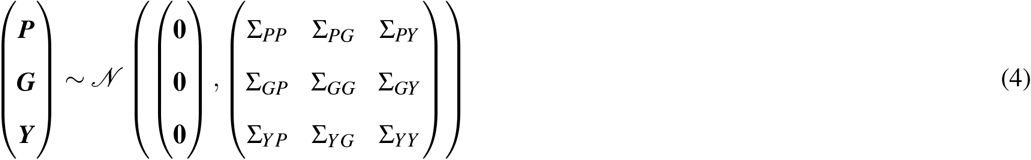

Based on the polygenic model assumptions, the covariance between the children’s genetic components and their parents’ corresponding genetic components is expected to be 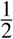, as each child inherits half of the trait-determining alleles from their mother, and the other half from their father. Hence, we specify empirical covariance matrices for Σ_*PP*_ and Σ_*GG*_:

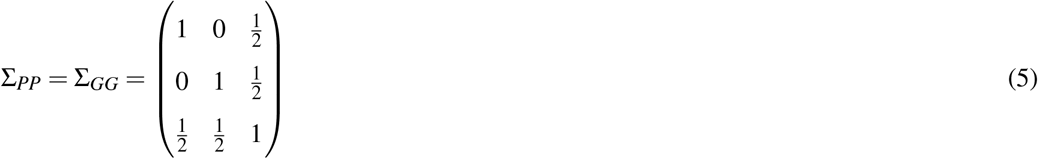

It is noteworthy that by specifying these variance-covariance matrices, we assume no consanguinity and no assortative mating.

Since ***P*** and ***G*** are assumed to be independent of each other, we set:

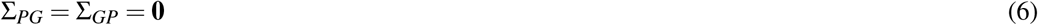

For associations between the trait and genetic components, we can derive that *Cov*(*P*_*k*_,*Y*_*k*_) = *Cov*(*P*_*k*_, *αP*_*k*_ + *β G*_*k*_ + *ε*_*k*_) = *α* for *k* ∈ {*M, F,C*} and 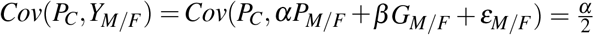 (the subscript *M/F* represents maternal or paternal component), thus

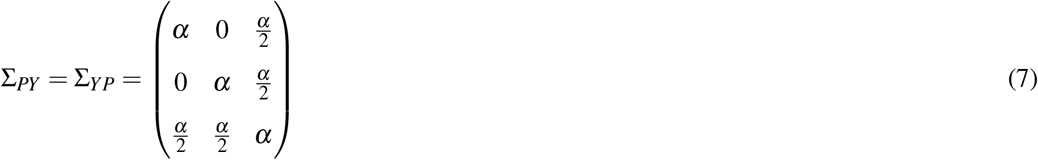

Similarly,

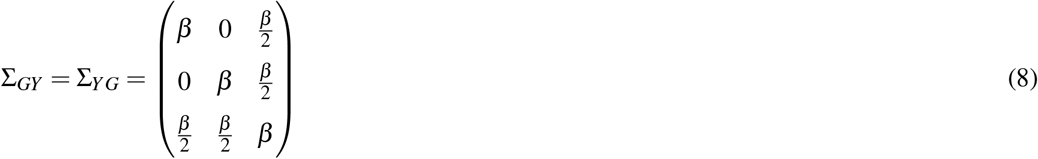

Furthermore, the covariance between the children’s trait and the parents’ trait 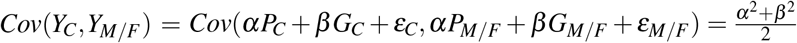. Therefore,

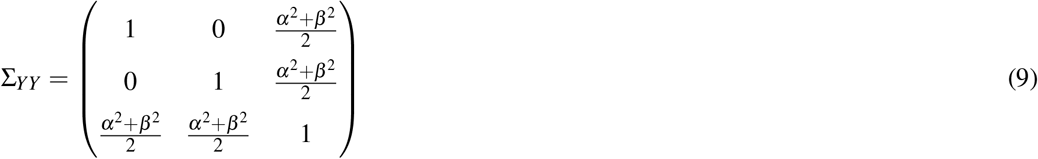

Equation 9 implies that if a combination of the parental trait measures, usually the mid-parental trait measure, is used to predict the children’s trait, the expected proportion of variance explained is

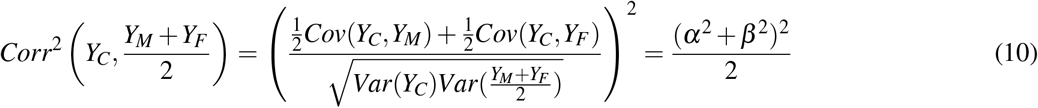

In practice, an estimate of *α* can be obtained from studies developing polygenic risk scores, i.e. 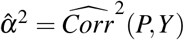. Meanwhile, observational studies reporting the magnitude of associations between parental trait measures and the children’s trait measures can inform the under-captured heritability *β*^2^, e.g.

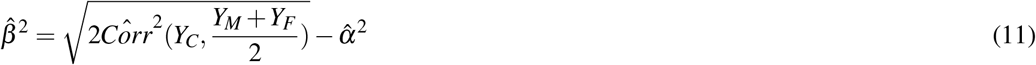

### Continuous trait prediction incorporating parental trait measures

Based on the distributional assumptions in Equation 4, we are able to infer *Y*_*C*_ with the parental trait measures (*Y*_*M*_ and/or *Y*_*F*_) and the children’s polygenic risk score (*P*_*C*_), if we have estimates of *α* and *β*. Specifically, given the properties of the multivariate normal distribution,

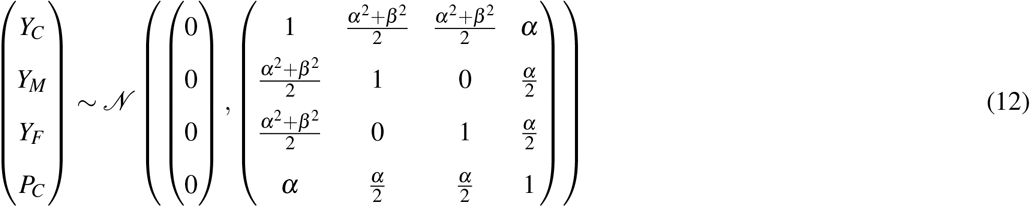

For the *j*-th individual with a polygenic risk score *p*_*C, j*_ and parental measures of the trait *y*_*M, j*_, *y*_*F, j*_, we use the conditional expectation as the predictor for the trait:

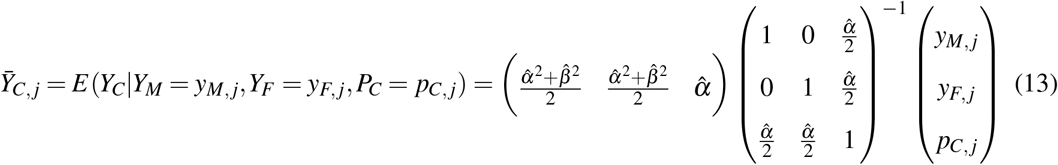

Alternatively, we can create a predictor using the polygenic risk score with one parental measure if only the maternal or the paternal measure is available:

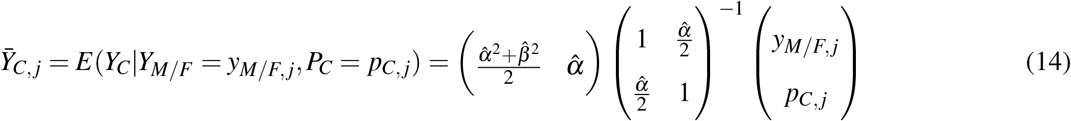

This framework is generalizable to any degree of family relationships, with modification of the variance-covariance matrices, possibly using empirical estimates of genetic relatedness. However, because information about more distantly related family members is more difficult to obtain, and is rarely complete in large cohort studies, and because parental information is the most relevant to risk prediction, we focus on parental trait measures or disease history in this work.

### Modelling latent genetic components for binary diseases

For binary diseases, we combine a liability model with the above latent factor model and explicitly model measurable covariate effects (such as age and sex) since they are non-trivial in this case. We denote the mothers’, fathers’ and children’s diseases of interest as *Z*_*M*_, *Z*_*F*_, and *Z*_*C*_, respectively. We assume that

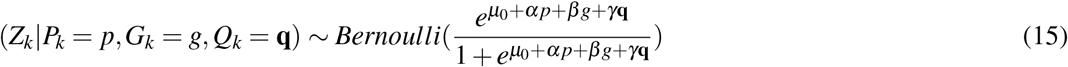

for *k* ∈ {*M, F,C*}, where *Q* represents measured (single or multiple) covariates with effects *γ*, and *μ*_0_ denotes the baseline odds (on the logarithmic scale) of the disease in the target population. *P* and *G* are assumed to be independently distributed with *P* ∼ *𝒩* (0, 1) and *G* ∼ *𝒩* (0, 1).

If we introduce *H* = *αP* + *β G*, where (*H* ∼ *𝒩* (0, *α*^2^ + *β*^2^)) includes both genetic components, then equation 15 simplifies to

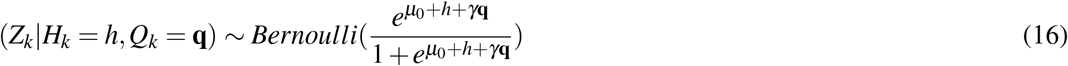

With the Bernoulli distribution assumptions, we adopt a logit-link in modelling the genetic liability, instead of the probit-link which more commonly used for liability threshold models^19^. This choice facilitates estimation of the model parameters from the results of widely-implemented logistic regression models. Nevertheless, heritability estimates can still be obtained. As shown previously^53^, an approximation to obtain the effect size estimate on the liability scale is

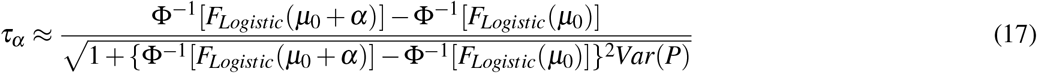

and

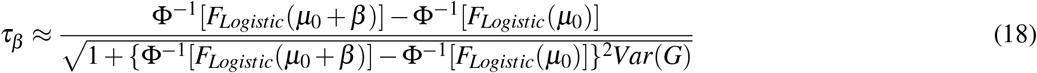

where Φ is the standard normal cumulative distribution function, *F*_*Logistic*_ is the cumulative distribution function of a logistic distribution with mean of 0 and variance of 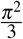, i.e. 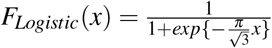, and *Var*(*P*) = *Var*(*G*) = 1.

Consequently, heritability captured by the polygenic risk score can be estimated as 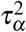, while the under-captured heritability can be estimated as 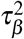. Notably, unlike the model for continuous traits (Section) where *α* and *β* are automatically bounded (0 ≤ *α*^2^ + *β*^2^ ≤ 1), here, the effects of the genetic components *P* and *G* are unconstrained.

Estimates of the unknown parameters can be obtained from summary statistics of existing observational studies. That is, we can directly obtain 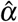 based on the magnitude of association between a polygenic risk score and the disease risk, i.e. odds ratio (OR) per one standard deviation increase in the polygenic risk score, together with 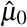 and 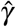. Furthermore, we can empirically estimate *β* if we have an estimate of the association between a parental disease history and the disease risk amongst the children.

Specifically, from the joint distribution

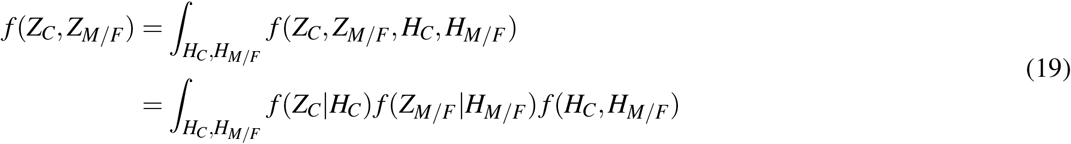

where

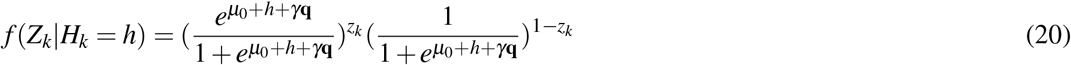

for *k* ∈ {*M, F,C*}, and

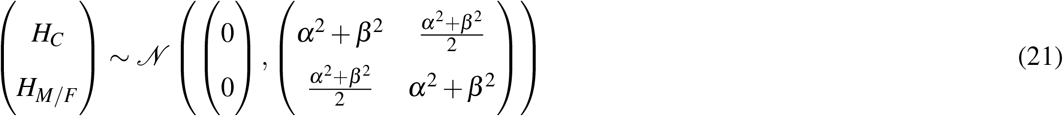

we have an explicit expression for the OR based on either the maternal or the paternal disease history as a function of the unknown *β* :

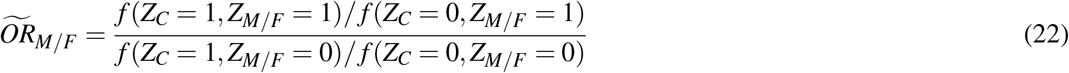

Therefore, we perform a numerical line search to obtain an empirical estimate of *β* such that the theoretical 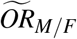 is close to the observed 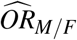:

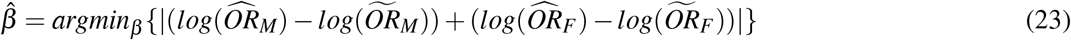

Note that the OR associated with the maternal disease history may differ from that based on the paternal disease history, because sex is included as a covariate (in *Q*) in this model.

### Binary disease risk prediction incorporating parental disease history

Similar to predicting a continuous trait (Section), we aim to infer *f* (*H*_*C*_|*Z*_*M*_, *Z*_*F*_, *P*_*C*_). since the distribution of *H*_*C*_ naturally informs *f* (*Z*_*C*_). Then we can use the conditional expectation as the predictor.

We observe that

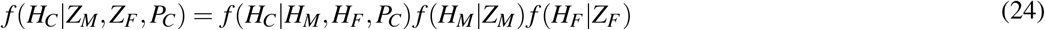

has no closed-form solution for its expectation, since

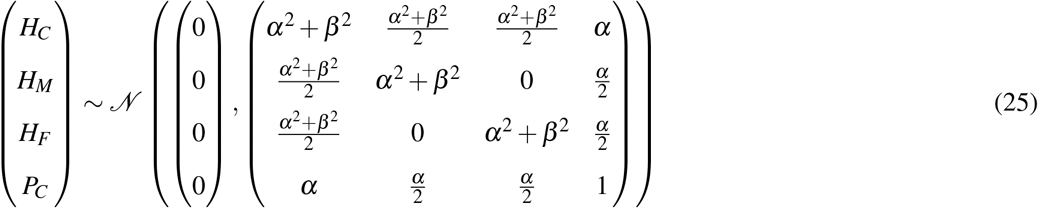

and

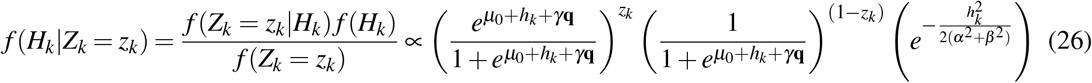

for *k* ∈ {*M, F,C*}. Therefore, we implement an importance sampling scheme to approximate this conditional distribution and derive its expectation:

- For the *j*-th individual with a polygenic risk score *p*_*C, j*_ and parental disease history *z*_*M, j*_ ∈ {0, 1}, *z*_*F, j*_ ∈ {0, 1}, we randomly generate *L* (a large number, e.g. 1,000,000) samples of *H*_*M, j*_ and *H*_*F, j*_ based on *f* (*H*_*M*_|*Z*_*M*_ = *z*_*M, j*_) and *f* (*H*_*F*_ |*Z*_*F*_ = *z*_*F, j*_) in Equation 26, respectively;
- For the *l*-th sample of *H*_*M, j*_ and *H*_*F, j*_, denoted as *h*_*M, j,l*_ and *h*_*F, j,l*_, we derive

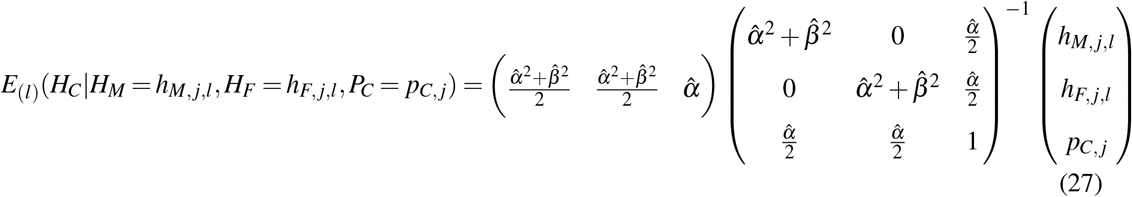
- We repeat for all *L* samples and obtain the predictor for the *j*-th individual as

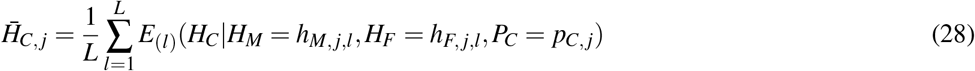

Alternatively, we can utilize the disease history of only one parent, particularly when a disease is highly sex-specific, where Equation 27 is modified as

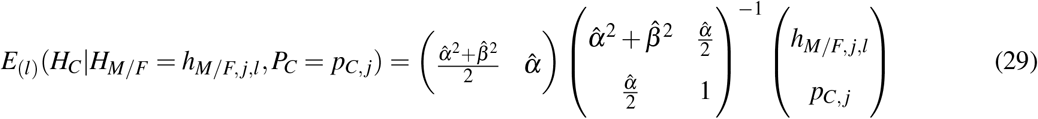

and Equation 28 is modified correspondingly as

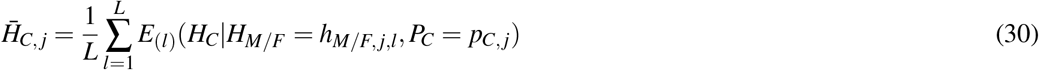

### Predicting adult height for children in the Avon Longitudinal Study of Parents and Children

From 1991 to 1992, the ALSPAC cohort recruited 14,541 pregnancies in the Bristol and Avon areas in the United Kingdom^28, 29^. In addition, 913 pregnancies were enrolled in later phases of the study. The total sample size for analyses using data collected after the age of seven is therefore 15,454 pregnancies, resulting in 15,589 foetuses. Of these, 14,901 were alive at 1 year of age. Ethical approval for the study was obtained from the ALSPAC Ethics and Law Committee and the Local Research Ethics Committees. Consent for biological samples has been collected in accordance with the Human Tissue Act (2004). Informed consent for the use of data collected via questionnaires and clinics was obtained from participants following the recommendations of the ALSPAC Ethics and Law Committee at the time.

Genotyping of the children initially recruited into this cohort was conducted using the Illumina HumanHap550 quad genotyping platforms. The genotypes were imputed to the 1000 Genomes Phase 3 reference panel^54^. Height of the biological parents was measured during clinical visits. Children’s adult height was measured at age 24. Measurement of height was performed using a Harpenden stadiometer (Holtain Ltd). Among the genotyped children of European ancestries who had measured adult standing height, 941 had both maternal and paternal height measures of biological parents, 1,305 only had maternal height measures of biological mothers, and 151 only had paternal height measures of biological fathers (**Supplementary Table 1**). We generated height z-scores by separately standardizing the children’s, mothers’ and fathers’ measured height to have zero mean and unit variance, where the standardization for the population of children was stratified by sex.

From studies that did not involve the ALSPAC cohort, we obtained an empirical estimate for *α* based on the association between a polygenic risk score for height z-score and the measured height z-score^7^. We further obtained an empirical estimate for *β* based on the association between the mid-parental height z-score and the measured height z-score^32^ based on Equation 11. Subsequently, following Section we derived predicted height z-score for each child by combining their calculated polygenic risk score with one or both parental height z-scores (Equation 13 for children with mid-parental height measures and Equation 14 for children with only maternal or paternal height measures).

We evaluated the proportion of variance explained and the RMSE (after transforming the predictor of z-scores back to the scale of absolute height measures) of the joint predictors, and compared these metrics of predictive performance with those based on the polygenic risk score, the parental height z-score alone.

Furthermore, a gold-standard predictor was obtained by fitting multivariate linear regression including both the polygenic risk score and parental height z-score as predictors on the test samples. This represents the upper-bound of predictive performance achievable by any linear predictors.

### Predicting risk of complex diseases in the UK Biobank

From 2006 to 2010, the UK Biobank study recruited approximately 500,000 participants who were aged between 40-69 years, at multiple recruitment centers in the United Kingdom^30^. Ethics approval for the UK Biobank study was obtained from the North West Centre for Research Ethics Committee (11/NW/0382). The UK Biobank ethics statement is available at https://www.ukbiobank.ac.uk/learn-more-about-uk-biobank/about-us/ethics. All UK Biobank participants provided informed consent at recruitment.

Upon recruitment, demographic and anthropometric information were collected. Genotyping of more than 480,000 participants were conducted using the Applied Biosystems™ UK BiLEVE Axiom™ Array or UK Biobank Axiom™ Array. The genotypes were imputed to the Haplotype Reference Consortium reference panel^55^.

Participants who had any of following complex diseases were identified based on inpatient International Classification of Diseases (ICD-10) diagnosis codes, Office of Population Censuses and Surveys (OPCS-4) procedure codes, or self-reported medical history during an interview with a trained nurse (**Supplementary Table 2**). ICD-10 codes for cancer diagnoses were retrieved by the UK Biobank through the national cancer registries. These disease outcomes included both prevalent cases identified upon initial recruitment and incident cases identified in more recent follow-up data collection.

- Breast cancer: ICD-10 code C50 (malignant neoplasm of breast), specific to women;
- Prostate cancer: ICD-10 code C61 (malignant neoplasm of prostate), specific to men;
- Colorectal cancer: ICD-10 codes C18 (malignant neoplasm of colon), C19 (malignant neoplasm of rectal sigmoid junction), or C20 (malignant neoplasm of rectum);
- Lung cancer: ICD-10 code C34 (malignant neoplasm of bronchus and lung);
- Myocardial infarction: ICD-10 code I21 (acute myocardial infarction);
- Ischemic heart disease: ICD-10 codes I20 (angina pectoris), I21 (acute myocardial infarction), I22 (subsequent myocardial infarction), I23 (complications following acute myocardial infarction), I24 (other acute ischemic heart diseases), or I25 (chronic ischemic heart disease), or OPCS-4 codes for coronary artery bypass grafting or coronary angioplasty with or without stenting;
- Stroke: ICD-10 codes I60 (subarachnoid haemorrhage), I61 (intracerebral haemorrhage), I62 (other nontraumatic intracranial haemorrhage), I63 (cerebral infarction), or I64 (stroke, not specified as haemorrhage or infarction);
- Type 2 diabetes: ICD-10 codes E11 (non-insulin-dependent diabetes mellitus), E13 (other specified diabetes mellitus), or E14 (unspecified diabetes mellitus), self-reported physician-made diagnosis, or self-reported use of anti-diabetic medications, excluding ICD-10 code E10 (insulin-dependent/type 1 diabetes mellitus);
- Alzheimer’s disease: ICD-10 code G30 (Alzheimer’s disease);
- Parkinson’s disease: ICD-10 code G20 (Parkinson’s disease);
- COPD: ICD-10 code J44 (chronic obstructive pulmonary disease), self-reported physician-made diagnosis, or self-reported use of medications for COPD.

Upon recruitment, a questionnaire inquired whether a participant had a parental history of breast cancer, prostate cancer, bowel cancer, lung cancer, heart disease, stroke, high blood pressure, diabetes, Alzheimer’s disease or dementia, Parkinson’s disease, or chronic bronchitis or emphysema. Further classifications of these diseases were not available, e.g. heart disease may include various types of diseases affecting the cardiovascular system, and diabetes may include both type 1 and type 2 diabetes. Participants who responded “do not know” or “prefer not to answer” were considered missing data. No participant reported paternal history of breast cancer or maternal history of prostate cancer. We matched the participants’ diseases with these parental records of diseases that had the same or a similar clinical definition.

Notably, while disease history of siblings was also available in the UK Biobank, it lacked information upon whether the sibling was a full- or half-sibling, and how many siblings were affected by the disease. Because these details were essential for correctly specifying the model, we refrained from incorporating sibling disease history in this study.

We retrieved polygenic risk scores for the above complex diseases from the PGS Catalog^33^. These polygenic risk scores were developed using different computational approaches based on source populations that did not overlap or at most slightly overlapped with the UK Biobank (**Supplementary Table 3**). We used the same well-powered polygenic risk score for predicting myocardial infarction and ischemic heart disease, as myocardial infarction is a manifestation of ischemic heart disease.

We first used a training dataset, comprising randomly selected 10% of the UK Biobank participants for deriving these parameters (**Supplementary Table 2** and **Supplementary Figure 1**). Specifically, for each disease, we fitted logistic regression models separately for the corresponding polygenic risk score and parental disease history (maternal disease history and paternal disease history as two independent variables), while including covariate effects of age, sex (except for breast cancer and prostate cancer), recruitment centre, genotyping array, and the first 10 genetic principal components. These two logistic regression models led to empirical estimates of 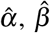, the baseline odds of disease 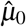, as well as the covariate effects 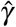 (Section ; **Supplementary Table 4**).

Next, we leveraged these parameters to obtain predictors of disease risk (at the liability scale; Section) for the rest 90% of the UK Biobank participants (**Supplementary Table 2** and **Supplementary Figure 1**). We evaluated the discriminative power of these joint predictors in identifying individuals at an elevated level of disease risk by AUROC and AUPRC. We tested whether the joint predictors could more accurately quantify the genetic risk than the polygenic risk score alone through comparing their AUROC by DeLong’s test^56^. We evaluated whether the risk stratification of the population could be improved by the joint predictors compared to the polygenic risk score by calculating NRI and IDI.

Last, we compared our joint predictors with a gold-standard numerical solution by jointly modelling the polygenic risk score and the parental disease history in a multivariate logistic regression model based on the training dataset (**Supplementary Figure 1**), and evaluating its discriminative power on the test dataset. To assess the robustness of this data-driven approach when individual-level data were insufficient, multivariate logistic regression models were fitted on randomly sampled subsets of the training dataset of smaller sample sizes (10%, 20%, and 50% of the original training dataset, each having 100 replicates). Predictive performance of these models was also evaluated on the test dataset.

We attempted to identify sources of the under-captured genetic component for Alzheimer’s disease and COPD, for which the joint predictors demonstrated the most improvements in risk stratification.

For Alzheimer’s disease, we determined the *APOE* genotype for each individual in the test dataset based on genotyping data of two SNPs: rs429358 and rs7412. We tested whether the polygenic risk score and the joint predictor were associated with carrying at least one e4 allele or carrying two e4 alleles, respectively. For COPD, we tested whether the polygenic risk score and the joint predictor were associated with self-reported smoking status (ever-smokers vs. never-smokers), respectively. All association tests were based on logistic regression, adjusted for the effects of age, sex, genotyping array, recruitment centre, and the first 10 genetic principal components.

## Data Availability

Individual genotype and pheontype data from the UK Biobank (https://www.ukbiobank.ac.uk/) and the ALSPAC (http://www.bristol.ac.uk/alspac/) are available through successful applications to the research committees. The ALSPAC website contains details of all the data that is available through a fully searchable data dictionary and variable search tool (http://www.bristol.ac.uk/alspac/researchers/our-data/). All other data are available from the corresponding author on reasonable request.

https://github.com/tianyuan-lu/PRS-FH-Prediction

## Code availability

All computational scripts for analyses in this study are available from the corresponding author on reasonable request. A computational toolkit implementing the latent factor model developed in this study is available at https://github.com/tianyuan-lu/PRS-FH-Prediction.

## Author Contributions

T.L. conceived and designed this study. T.L. and C.M.T.G. developed methodology. T.L. created software package. V.F. and T.L. acquired and managed data. T.L. performed analyses and visualized results. T.L. interpreted results with C.M.T.G., J.B.R. and V.F. C.M.T.G. supervised this study. T.L. drafted the manuscript. All authors read and revised the manuscript.

## Competing Interests

The authors declare the following competing interests: J.B.R. is the founder of 5 Prime Sciences, and has served as a consultant to GlaxoSmithKline and Deerfield Capital for their genetics programs. The other authors have no relevant disclosures.

## Acknowledgements

This research has been conducted using the UK Biobank resource under Application Number 27449 and 60755, and the ALSPAC resource under Application Number B3359. ALSPAC children were genotyped using the Illumina HumanHap550 quad chip genotyping platforms by Sample Logistics and Genotyping Facilities at Wellcome Sanger Institute and LabCorp (Laboratory Corporation of America), using support from 23andMe. We thank all the families who took part in the ALSPAC study, the midwives for their help in recruiting them, and the whole ALSPAC team, which includes interviewers, computer and laboratory technicians, clerical workers, research scientists, volunteers, managers, receptionists, and nurses. This study was enabled in part by support provided by Calcul Québec and Compute Canada. T.L. thanks Wenmin Zhang for helpful discussion about the latent factor models and prediction schemes.

C.M.T.G. is supported by a Canadian Institutes of Health Research grant (CIHR; PJT-148620). The J.B.R. research group is supported by the Canadian Institutes of Health Research (365825; 409511), the Lady Davis Institute of the Jewish General Hospital, the Canadian Foundation for Innovation, the NIH Foundation, Cancer Research UK, Genome Québec, the Public Health Agency of Canada and the Fonds de Recherche Québec Santé (FRQS). J.B.R. is supported by a FRQS Clinical Research Scholarship Merite. T.L. has been supported by a Vanier Canada Graduate Scholarship, an FRQS Doctoral Training Fellowship and a McGill University Faculty of Medicine Scholarship. The UK Medical Research Council and Wellcome (217065/Z/19/Z) and the University of Bristol provide core support for ALSPAC. A comprehensive list of grants funding is available on the ALSPAC website (http://www.bristol.ac.uk/alspac/external/documents/grant-acknowledgements.pdf). This research was specifically funded by Wellcome Trust and the Medical Research Council (076467/Z/05/Z).

## Notes

### Funding Statement

C.M.T.G. is supported by a Canadian Institutes of Health Research grant (CIHR; PJT-148620). The Richards research group is supported by the Canadian Institutes of Health Research (365825; 409511), the Lady Davis Institute of the Jewish General Hospital, the Canadian Foundation for Innovation, the NIH Foundation, Cancer Research UK, Genome Quebec, the Public Health Agency of Canada and the Fonds de Recherche Quebec Sante (FRQS). J.B.R. is supported by a FRQS Clinical Research Scholarship Merite. T.L. has been supported by a Vanier Canada Graduate Scholarship, an FRQS Doctoral Training Fellowship and a McGill University Faculty of Medicine Scholarship. The UK Medical Research Council and Wellcome (217065/Z/19/Z) and the University of Bristol provide core support for ALSPAC. A comprehensive list of grants funding is available on the ALSPAC website (http://www.bristol.ac.uk/alspac/external/documents/grant-acknowledgements.pdf). This research was specifically funded by Wellcome Trust and the Medical Research Council (076467/Z/05/Z).

### Author Declarations

IRB of the Lady Davis Institute for Medical Research at Montreal's Jewish General Hospital gave ethical approval for this work.

