## Supplementary Materials for "Capturing additional genetic risk from family history for improved polygenic risk prediction"

#### Supplementary Note 1: Simulation design

As a proof of concept, we conducted simulation studies to evaluate the utility and robustness of the proposed method.
Based on the data generative process described in **Methods**, we first simulated genetic components $P$ and $G$ for $N = 20,000$ child-parent trios, respectively.

$$\begin{pmatrix} \boldsymbol{P} \\ \boldsymbol{G} \end{pmatrix} \sim \mathcal{N}\left(\begin{pmatrix} \boldsymbol{0} \\ \boldsymbol{0} \end{pmatrix}, \begin{pmatrix} \Sigma_{PP} & \Sigma_{PG} \\ \Sigma_{GP} & \Sigma_{GG} \end{pmatrix}\right) \tag{1}$$

where $\boldsymbol{P} = (P_M, P_F, P_C)^\top$, $\boldsymbol{G} = (G_M, G_F, G_C)^\top$, and

$$\Sigma_{PP} = \Sigma_{GG} = \begin{pmatrix} 1 & 0 & \frac{1}{2} \\ 0 & 1 & \frac{1}{2} \\ \frac{1}{2} & \frac{1}{2} & 1 \end{pmatrix} \tag{2}$$

For each mother, father, or child, a continuous trait was generated as

$$Y_{k,j} = \alpha P_{k,j} + \beta G_{k,j} + \epsilon_{k,j} \tag{3}$$

where $\epsilon_{k,j}$ was sampled from $\mathcal{N}(0, 1 - (\alpha^2 + \beta^2))$, for $k \in \{M, F, C\}$ and $j \in \{1, ..., N\}$.
Then, a binary outcome was generated as

$$Z_{k,j} \sim Bernoulli(\frac{e^{\mu_0 + Y_{k,j}}}{1 + e^{\mu_0 + Y_{k,j}}}) \tag{4}$$

for $k \in \{M, F, C\}$ and $j \in \{1, ..., N\}$, with $\mu_0 = -1$.
We randomly selected 10,000 child-parent trios into a training (reference) dataset to derive model parameters (**Methods**). For the continuous trait, a joint predictor would combine $P_C$, $Y_M$ and $Y_F$; For the binary outcome, a joint predictor would combine $P_C$, $Z_M$ and $Z_F$. The remaining 10,000 child-parent trios constituted a test dataset, where the joint predictor (FHPRS) was compared to $P_C$ (i.e. prediction using the polygenic risk score, PRS, alone) in predicting $Y_C$ or $Z_C$.
For the continuous trait, we evaluated the increment in the proportion of variance explained

$$\Delta_{metric,continuous} = R^2_{FHPRS} - R^2_{PRS} \tag{5}$$

For the binary trait, we evaluated the increment in the area under the receiver operating characteristic curve (AUROC)

$$\Delta_{metric,binary} = AUROC_{FHPRS} - AUROC_{PRS} \tag{6}$$

We experimented combinations of $\alpha$ ($\alpha \in \{0.1, 0.2, 0.5\}$) and $\beta$ ($\beta \in \{0.1, 0.2, 0.5\}$) to assess model performance under different magnitudes of genetic effects. We repeated each combination of $\alpha$ and $\beta$ 100 times.

As expected, the joint predictor mostly outperformed predictions using the polygenic risk score alone for both continuous and binary outcomes when the proportion of under-modelled variance was moderate or large ($\beta = 0.2$ or $\beta = 0.5$), although it may have worse performance with $\beta = 0.1$ due to excessive random errors (**Supplementary Figure 4**). Not surprisingly, improvements of AUROC were marginal even in settings of a high $\beta$ (**Supplementary Figure 4**), reflecting that the metric of AUROC is not very sensitive to addition of meaningful predictors [1].

#### Supplementary Note 2: Errors in model parameter estimation

Next, we explored to what extent our method could tolerate errors in model parameter estimation as precise estimates were usually elusive in real data. Specifically, we provided inaccurate $\alpha$ and $\beta$ as input parameters

$$\tilde{\alpha} = \delta_\alpha \alpha \tag{7}$$

$$\tilde{\beta} = \delta_\beta \beta \tag{8}$$

where $\alpha$ and $\beta$ were true model parameters in each simulation setting, with error factors $\delta_\alpha$ and $\delta_\beta$ taking value in $\{0.5, 0.8, 0.9, 1.1, 1.2, 1.5\}$.
We focused on the continuous trait in this analysis and derived $\Delta_{metric,continuous}$ as described in Equation 5, for each of the 100 repeats of each simulation setting.
We found that the joint predictor was robust to slight-to-moderate errors in model parameter estimation (**Supplementary Figures 5-7**). When the under-modelled genetic component was weak ($\beta = 0.1$; **Supplementary Figure 5**), as expected, the joint predictors did not demonstrate notable improvements over the polygenic risk scores even if the model parameters were estimated accurately. When the under-modelled genetic component was moderate or strong ($\beta = 0.2$ or $\beta = 0.5$), the model performance remained largely unchanged with error factors between 0.9 and 1.1 (**Supplementary Figures 6** and **7**). Moreover, the joint predictors maintained improvements over polygenic risk scores with error factors between 0.8 and 1.2, though the magnitudes of improvements may attenuate (**Supplementary Figures 6** and **7**). In contrast, when the model parameters were severely under- or over-estimated by 50%, the joint predictors could have worse predictive performance than using the polygenic risk scores alone (**Supplementary Figures 6** and **7**).

#### Supplementary Note 3: Violations of model assumptions

Furthermore, we evaluated whether our method could withstand possible violations of model assumptions. For this analysis, we fixed $\alpha = 0.5$ and $\beta = 0.5$ in Equation 5.

##### • Assortative mating

When assortative mating occurs, the independence assumption upon maternal and paternal components would be violated. We explored three scenarios below with a parameter $\rho$ to quantify the intensity of assortative mating.

###### Assotative mating driven by non-genetic factors

Namely, the residual variance components in Equation 3 could be correlated

$$\begin{pmatrix}\epsilon_M \\ \epsilon_F \\ \epsilon_C\end{pmatrix} \sim \mathcal{N}\left(\begin{pmatrix}0\\0\\0\end{pmatrix}, \begin{pmatrix}1-(\alpha^2+\beta^2) & \rho(1-(\alpha^2+\beta^2)) & 0 \\ \rho(1-(\alpha^2+\beta^2)) & 1-(\alpha^2+\beta^2) & 0 \\ 0 & 0 & 1-(\alpha^2+\beta^2)\end{pmatrix}\right) \tag{9}$$

###### Assotative mating driven by under-modelled genetic component

Namely,

$$\Sigma_{GG} = \begin{pmatrix} 1 & \rho & \frac{1}{2} \\ \rho & 1 & \frac{1}{2} \\ \frac{1}{2} & \frac{1}{2} & 1 \end{pmatrix} \tag{10}$$

###### Assotative mating driven by genetic component captured by the polygenic risk score

Namely,

$$\Sigma_{PP} = \begin{pmatrix} 1 & \rho & \frac{1}{2} \\ \rho & 1 & \frac{1}{2} \\ \frac{1}{2} & \frac{1}{2} & 1 \end{pmatrix} \tag{11}$$

In all three scenarios, we experimented with $\rho \in \{0.1, 0.2, 0.5\}$ and derived $\Delta_{metric,continuous}$ as described in Equation 5, for each of the 100 repeats of each simulation setting.
Encouragingly, while assortative mating did lead to attenuation of model performance and the impact increased with larger $\rho$, the joint predictors could still consistently outperform predictions using polygenic risk scores alone (**Supplementary Figure 8**).

##### • Interaction effects

Modelling interaction effects is non-trivial in the current model design. Again, we explored three scenarios of gene-by-environment and gene-by-gene interactions to evaluate the robustness of our method, with a parameter $\gamma$ to quantify the magnitude of interaction effect.

###### Interaction between under-modelled genetic component and non-genetic factors

Here, Equation 3 was modified as

$$Y_{k,j} = \alpha P_{k,j} + \beta G_{k,j} + \epsilon_{k,j} + \gamma G_{k,j}\epsilon_{k,j} \tag{12}$$

for $k \in \{M, F, C\}$ and $j \in \{1, ..., N\}$.

###### Interaction between modelled genetic component in the polygenic risk score and non-genetic factors

Here, Equation 3 was modified as

$$Y_{k,j} = \alpha P_{k,j} + \beta G_{k,j} + \epsilon_{k,j} + \gamma P_{k,j}\epsilon_{k,j} \tag{13}$$

for $k \in \{M, F, C\}$ and $j \in \{1, ..., N\}$.

###### Interaction between under-modelled and modelled genetic components

Here, Equation 3 was modified as

$$Y_{k,j} = \alpha P_{k,j} + \beta G_{k,j} + \epsilon_{k,j} + \gamma P_{k,j} G_{k,j} \tag{14}$$

for $k \in \{M, F, C\}$ and $j \in \{1, ..., N\}$.
In all three scenarios, we experimented with $\gamma \in \{0.1, 0.2, 0.5\}$ and derived $\Delta_{metric,continuous}$ as described in Equation 5, for each of the 100 repeats of each simulation setting.
As expected, in the presence of gene-by-environment interactions, the joint predictors would have worse performance than in simulation settings without these interaction effects. However, again, they

could still consistently outperform predictions using polygenic risk scores alone (**Supplementary Figure 9**).
On the contrary, when gene-by-gene interaction effects existed, the joint predictors demonstrated more prominent improvements over polygenic risk scores as the interaction effects became stronger. This was because the overall trait variance attributable to genetics increased and our model incorporated both genetic components (**Supplementary Figure 9**).

- **Non-normal distribution of $G$**

Last, the under-modelled genetic component $G$ may contain rare pathogenic variants having a large effect. Such genetic effects are unlikely to follow a normal distribution as specified in the current model design.
We suppose there are $\kappa$ independent loci in the populations of mothers and fathers, respectively. We further suppose that each effective allele of any of these $\kappa$ loci increases the phenotypic value $Y$ by 1 unit and that all effective allele carriers are heterozygotes due to the rarity and strong effect of these variants. The total number of large-effect alleles that a mother or a father carries hence follows $G_{M/F} \sim Binomial(\kappa, f)$, where $f$ is the overall frequency of rare variants. Therefore,

$$Var(G) = \kappa f(1 - f) \tag{15}$$

In the population of children, each child would inherit one allele from their mother and one from their father, for each of the $\kappa$ loci. Thus, the total number of large-effect alleles that the $j$-th child carries is generated by

$$G_{C,j} \sim Binomial(G_{M,j}, 0.5) + Binomial(G_{F,j}, 0.5) \tag{16}$$

for $j \in \{1, ..., N\}$.
We experimented with $\kappa \in \{2, 4, 6\}$ and derived $\Delta_{metric,continuous}$ as described in Equation 5, for each of the 100 repeats of each simulation setting. In each simulation setting, we calculated $f = \frac{1-\sqrt{1-\frac{4\beta^2}{\kappa}}}{2}$ accordingly.
As a result, the non-normal distribution of $G$ did not lead to substantial decrease of model performance. Instead, the degree of performance improvement achieved by the joint predictors largely overlapped with that obtained under simulation settings with normally distributed $G$ (**Supplementary Figure 10**).

### Summary

In summary, these simulation studies suggest that a joint predictor is able to consistently improve polygenic risk prediction by incorporating family history information. Importantly, the proposed model is not sensitive to errors in model parameter estimation and can tolerate slight-to-moderate violations of several model assumptions.

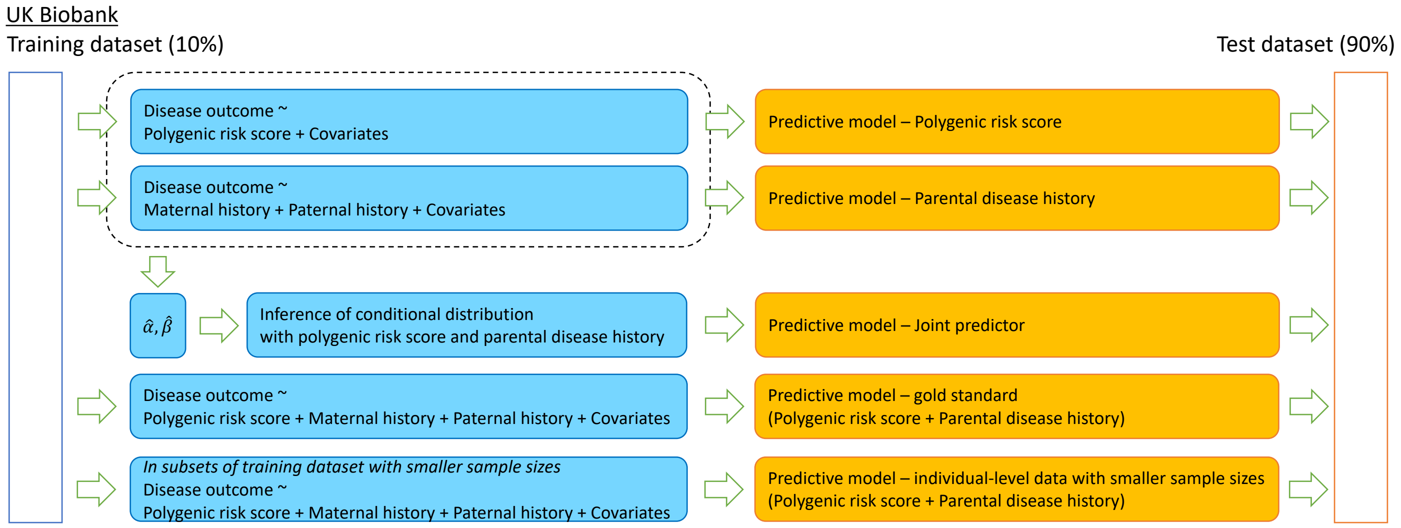


Figure S1. Development and evaluation of predictors in the UK Biobank. The UK Biobank was randomly split into a training dataset comprising 10% of the samples and a test dataset comprising 90% of the samples. For each disease, on the test dataset, the magnitude of association between the polygenic risk score as well as the parental disease history and the disease risk was estimated. These estimates were utilized to obtain a joint predictor with the latent factor model. A multivariate logistic regression model including the polygenic risk score and the parental disease history was fitted to obtain the gold-standard predictor. Furthermore, 10%, 20%, or 50% of the training dataset was randomly selected and used to fit multivariate logistic regression models, each repeating 100 times. These generate data-driven predictors based on individual-level data of smaller sample sizes. All predictors were tested on the test dataset.

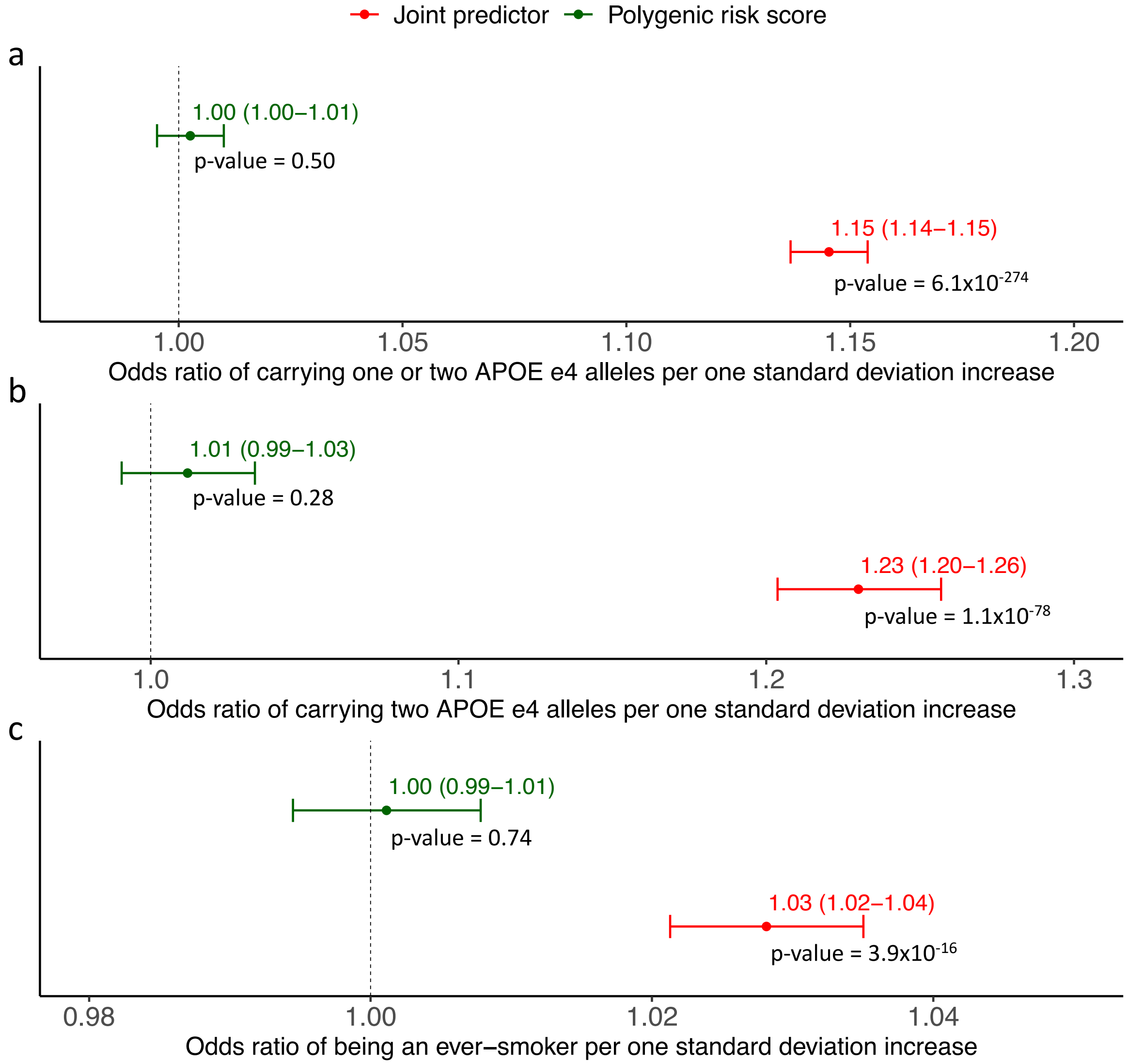


Figure S2. Association between genetic risk predictors and known risk factors. (a) Associations between genetic risk predictors for Alzheimer's disease and carrying at least one *APOE* e4 allele. (b) Associations between genetic risk predictors for Alzheimer's disease and carrying two *APOE* e4 alleles. In the UK Biobank test dataset for Alzheimer's disease, there were 8,616 e4/e4 homozygotes, 85,613 e3/e4 heterozygotes, 209,466 e3/e3 homozygotes, 44,293 e2/e3 heterozygotes, and 2,280 e2/e2 homozygotes. In addition, 9,113 individuals had ambiguous genotypes due to heterozygosity at both the rs429358 and rs7412 SNPs, and were not included in these association tests. (c) Associations between genetic risk predictors for COPD and being ever-smokers. There were 161,940 ever-smokers and 196,081 never-smokers. All association tests were based on individuals in the test dataset, adjusted for the effects of age, sex, genotyping array, recruitment center, and the first 10 genetic principal components. Error bars indicate 95% confidence intervals for the estimated odds ratios.

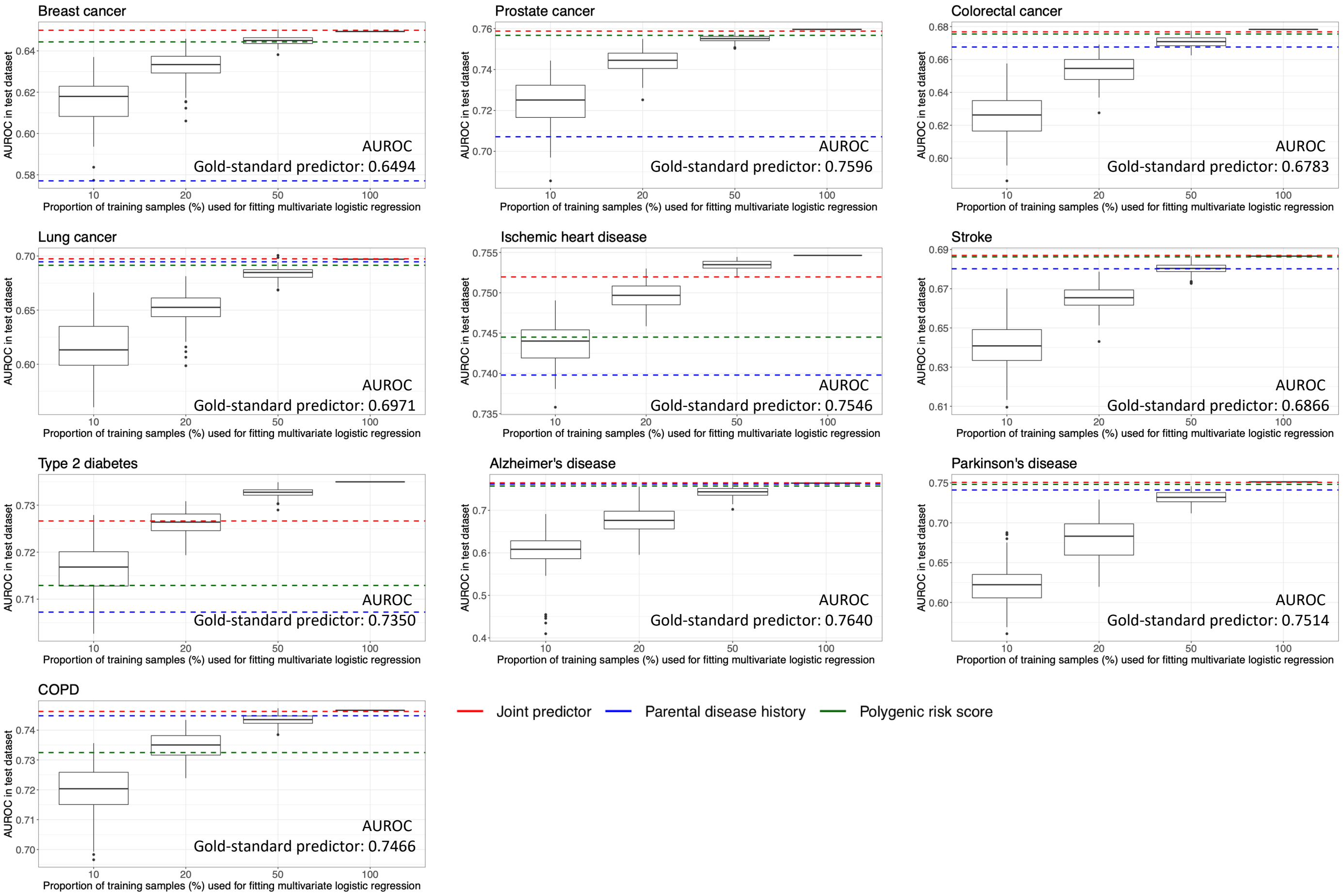


Figure S3. Performance of data-driven joint predictors obtained from multivariate logistic regression for 10 complex diseases. AUROC of the corresponding joint predictor, polygenic risk score, and parental disease history are summarized in Table 1, and are indicated by red, green, and blue dashed lines, respectively. Apart from ischemic heart disease and type 2 diabetes, the joint predictors obtained from the latent factor model have comparable discriminative power to the gold-standard predictors obtained from multivariate logistic regression based on all samples in the training dataset, and the joint predictors outperform other data-driven predictors based on subsets of the training dataset of smaller sample sizes. Box plots represent distributions of AUROC obtained in 100 replicates corresponding to different sample sizes.

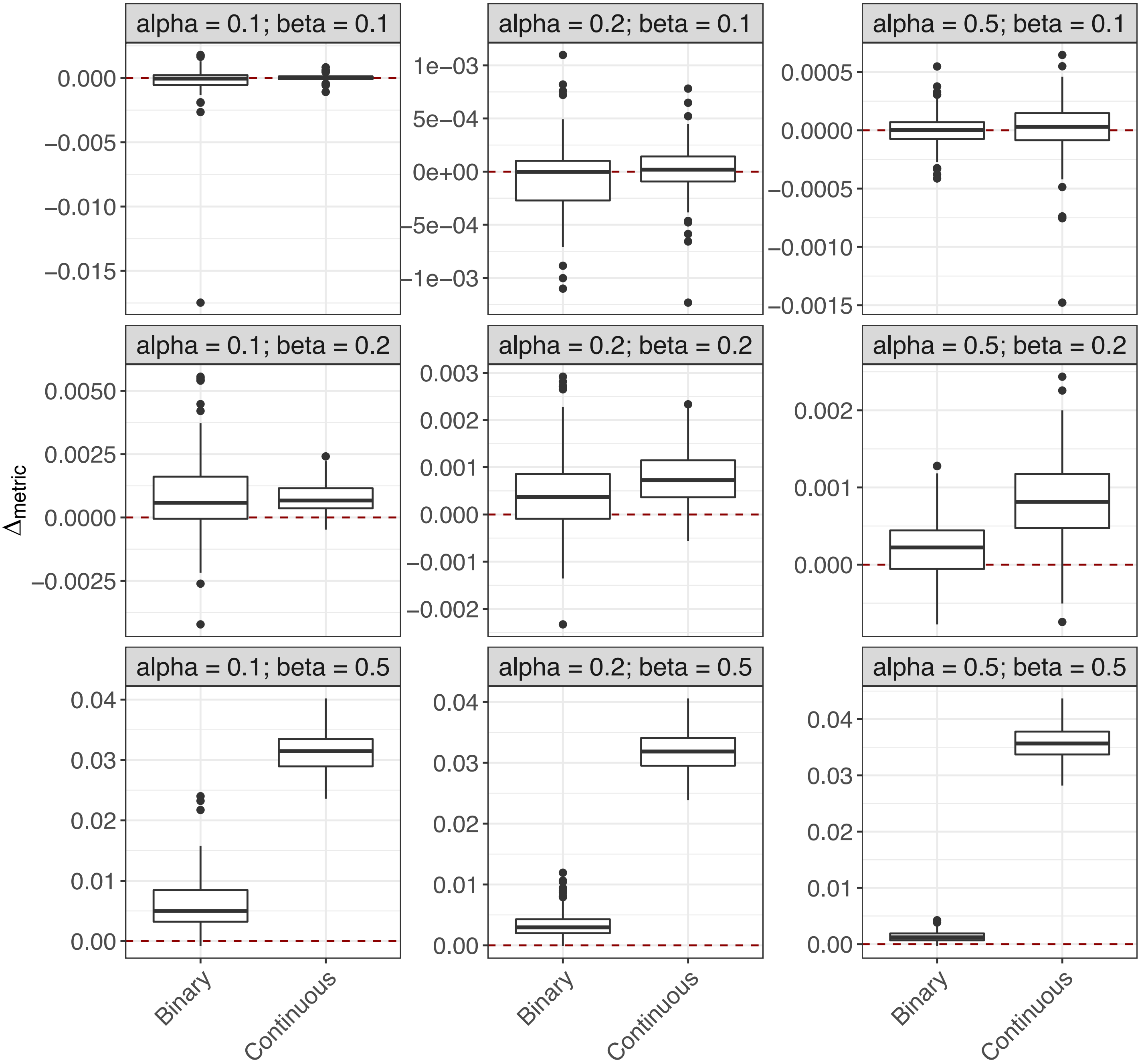


Figure S4. Comparison of performance between a joint predictor and polygenic risk score. Model parameters for each simulation setting are indicated. $\Delta_{metric}$ was defined as difference in proportion of variance explained for the simulated continuous trait and difference in area under the receiver operating characteristic curve for the simulated binary outcome based on the test dataset. A positive $\Delta_{metric}$ indicated that the joint predictor had improved predictive performance compared to the polygenic risk score. All simulation settings were repeated 100 times to obtain the distributions of $\Delta_{metric}$, represented by box plots.

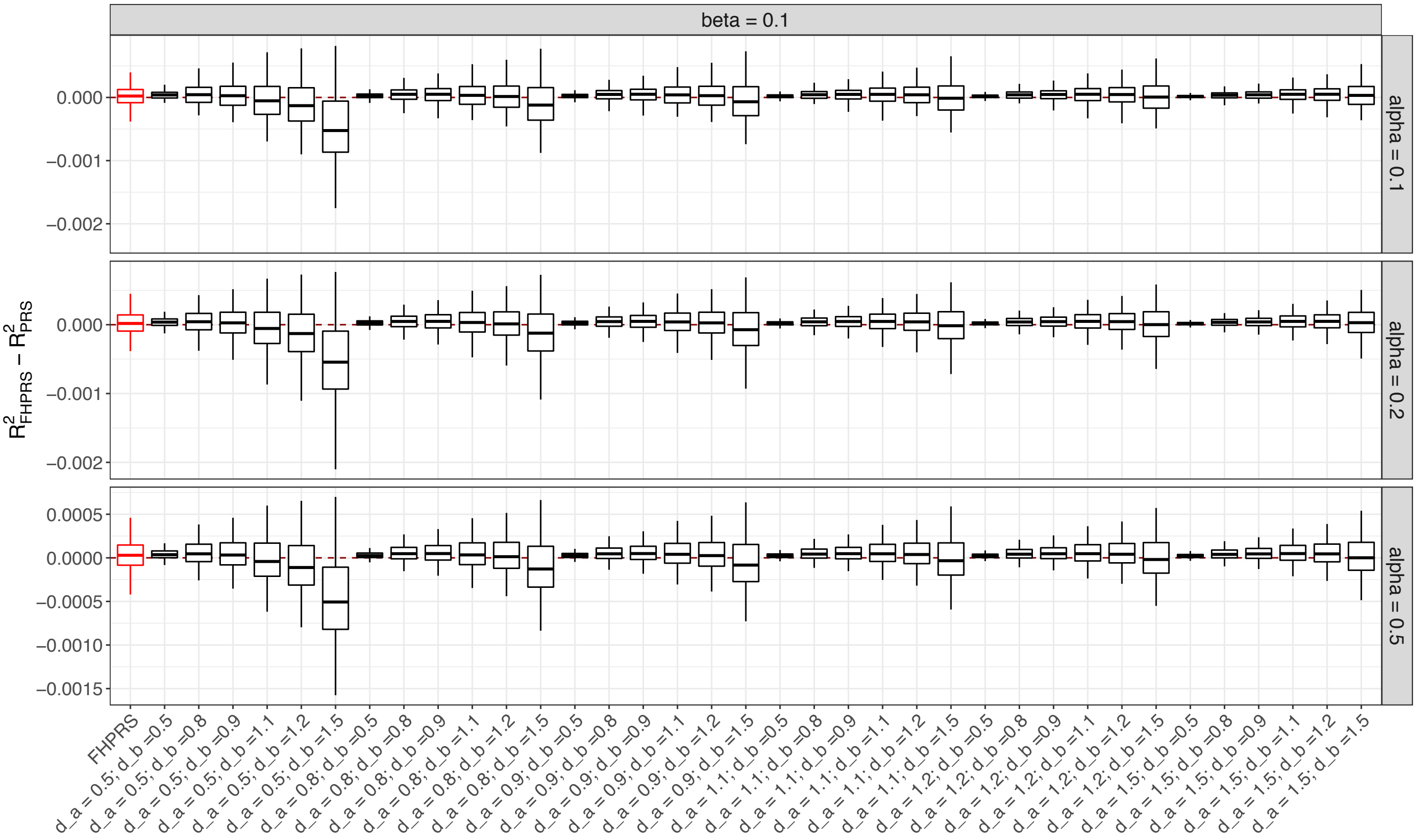


Figure S5. Impact of error in parameter estimation with a weak under-modelled genetic component ($\beta = 0.1$). The difference in proportion of variance explained for the simulated continuous trait based on the test dataset was calculated under different degrees of parameter estimation error, specified by error factors $\delta_\alpha$ and $\delta_\beta$ (Supplementary Notes). All simulation settings were repeated 100 times to obtain the distributions of the difference in proportion of variance explained, represented by box plots. Results derived from estimated parameters based on the training dataset are colored red. FHPRS: family history-assisted polygenic risk score; PRS: polygenic risk score.

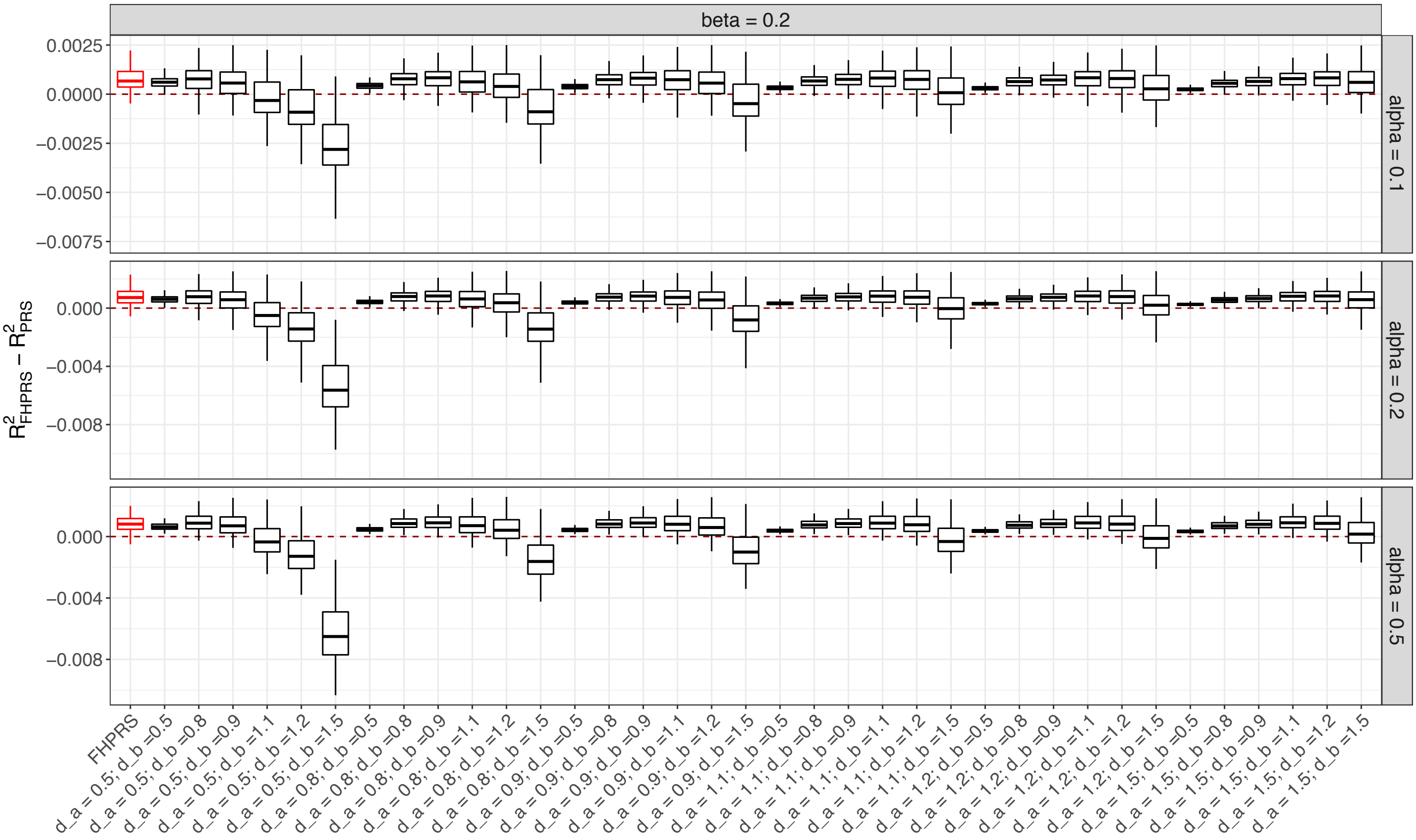


Figure S6. Impact of error in parameter estimation with a moderate under-modelled genetic component ($\beta = 0.2$). The difference in proportion of variance explained for the simulated continuous trait based on the test dataset was calculated under different degrees of parameter estimation error, specified by error factors $\delta_\alpha$ and $\delta_\beta$ (Supplementary Notes). All simulation settings were repeated 100 times to obtain the distributions of the difference in proportion of variance explained, represented by box plots. Results derived from estimated parameters based on the training dataset are colored red. FHPRS: family history-assisted polygenic risk score; PRS: polygenic risk score.

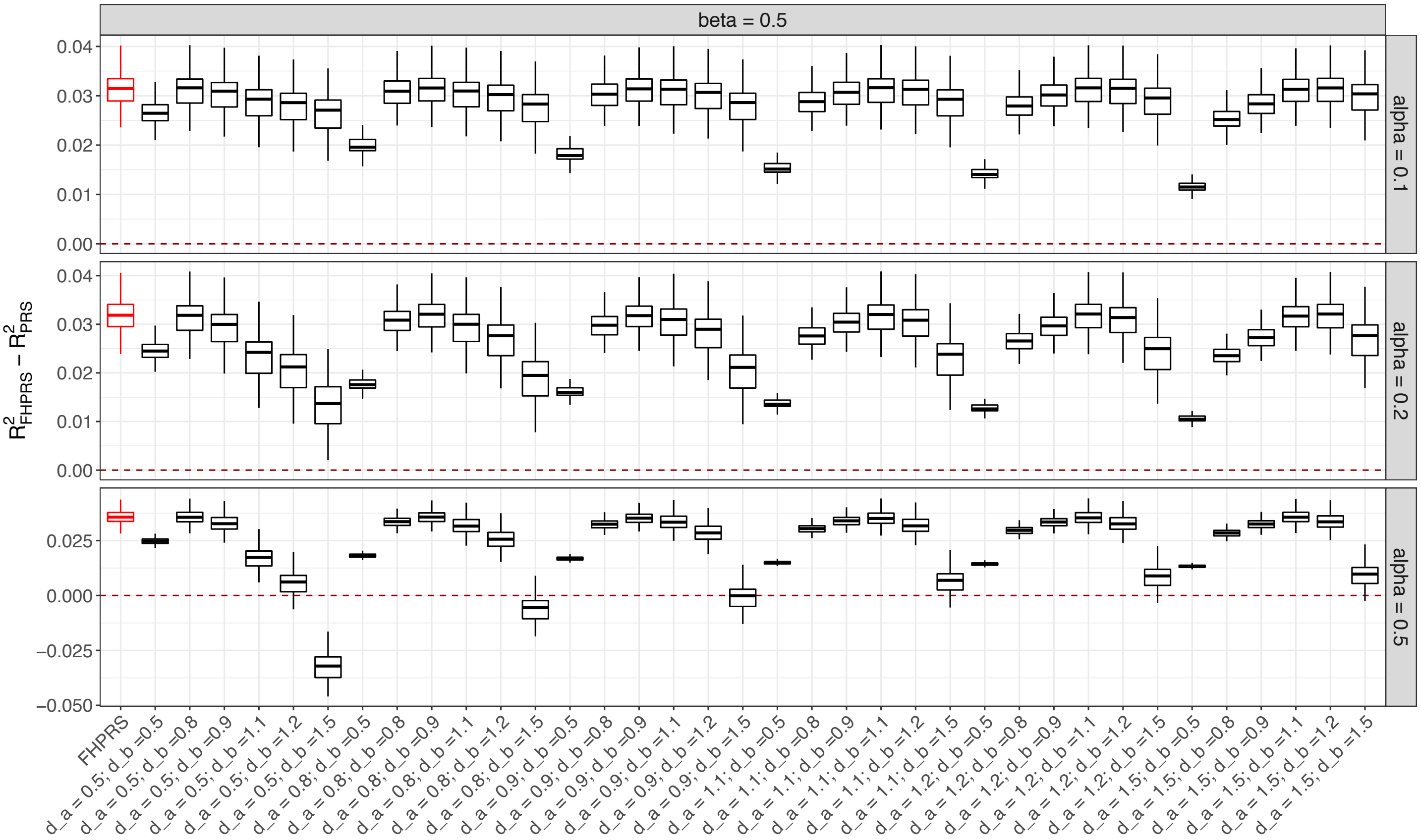


Figure S7. Impact of error in parameter estimation with a strong under-modelled genetic component ($\beta = 0.5$). The difference in proportion of variance explained for the simulated continuous trait based on the test dataset was calculated under different degrees of parameter estimation error, specified by error factors $\delta_\alpha$ and $\delta_\beta$ (Supplementary Notes). All simulation settings were repeated 100 times to obtain the distributions of the difference in proportion of variance explained, represented by box plots. Results derived from estimated parameters based on the training dataset are colored red. FHPRS: family history-assisted polygenic risk score; PRS: polygenic risk score.

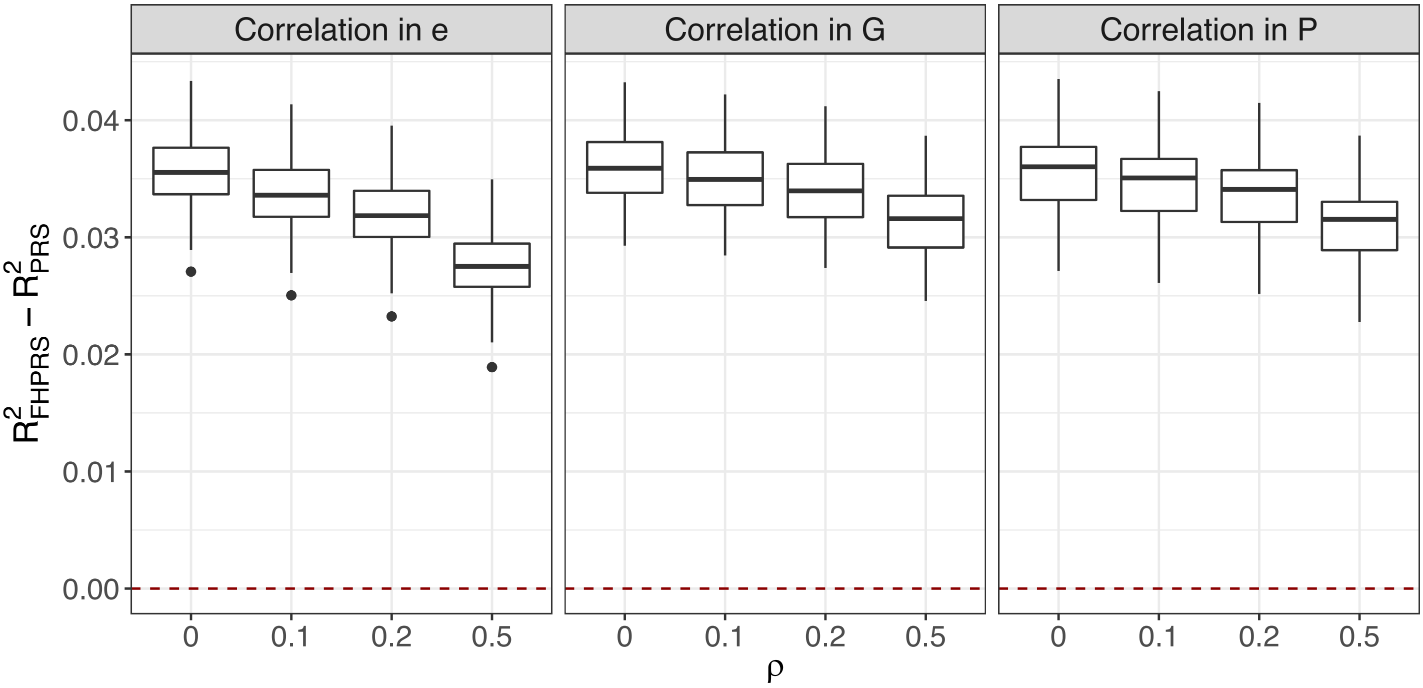


Figure S8. Impact of assortative mating on model performance. The difference in proportion of variance explained for the simulated continuous trait based on the test dataset was calculated in the presence of assortative mating driven by non-genetic factors (e), the under-modelled genetic component (G), or the modelled genetic component (P), respectively (Supplementary Notes). $\rho = 0$ represents simulation settings without assortative mating. All simulation settings were repeated 100 times with $\alpha = 0.5$ and $\beta = 0.5$ to obtain the distributions of the difference in proportion of variance explained, represented by box plots. FHPRS: family history-assisted polygenic risk score; PRS: polygenic risk score.

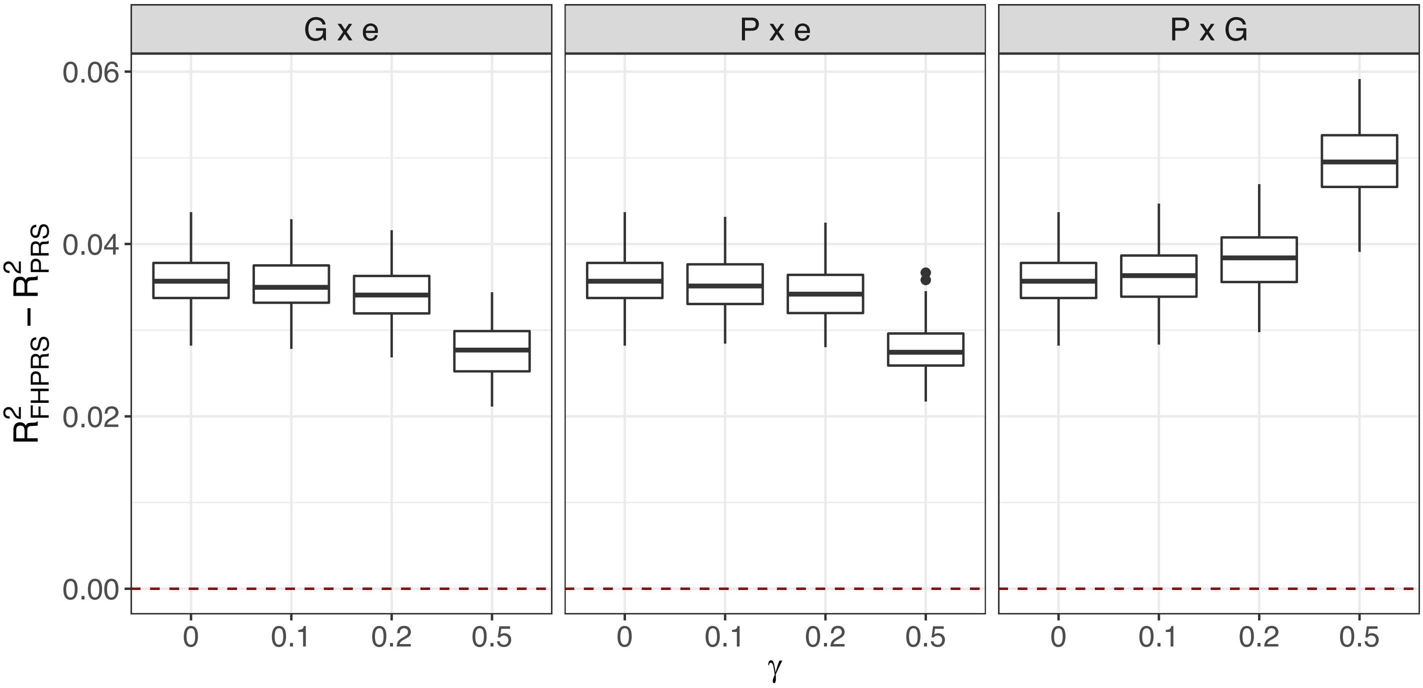


Figure S9. Impact of interaction effects on model performance. The difference in proportion of variance explained for the simulated continuous trait based on the test dataset was calculated in the presence of interaction between under-modelled genetic component and environmental factors (G x e), modelled genetic component and environmental factors (P x e), and the two genetic components (P x G), respectively (Supplementary Notes). $\gamma = 0$ represents simulation settings without interaction effects. All simulation settings were repeated 100 times with $\alpha = 0.5$ and $\beta = 0.5$ to obtain the distributions of the difference in proportion of variance explained, represented by box plots. FHPRS: family history-assisted polygenic risk score; PRS: polygenic risk score.

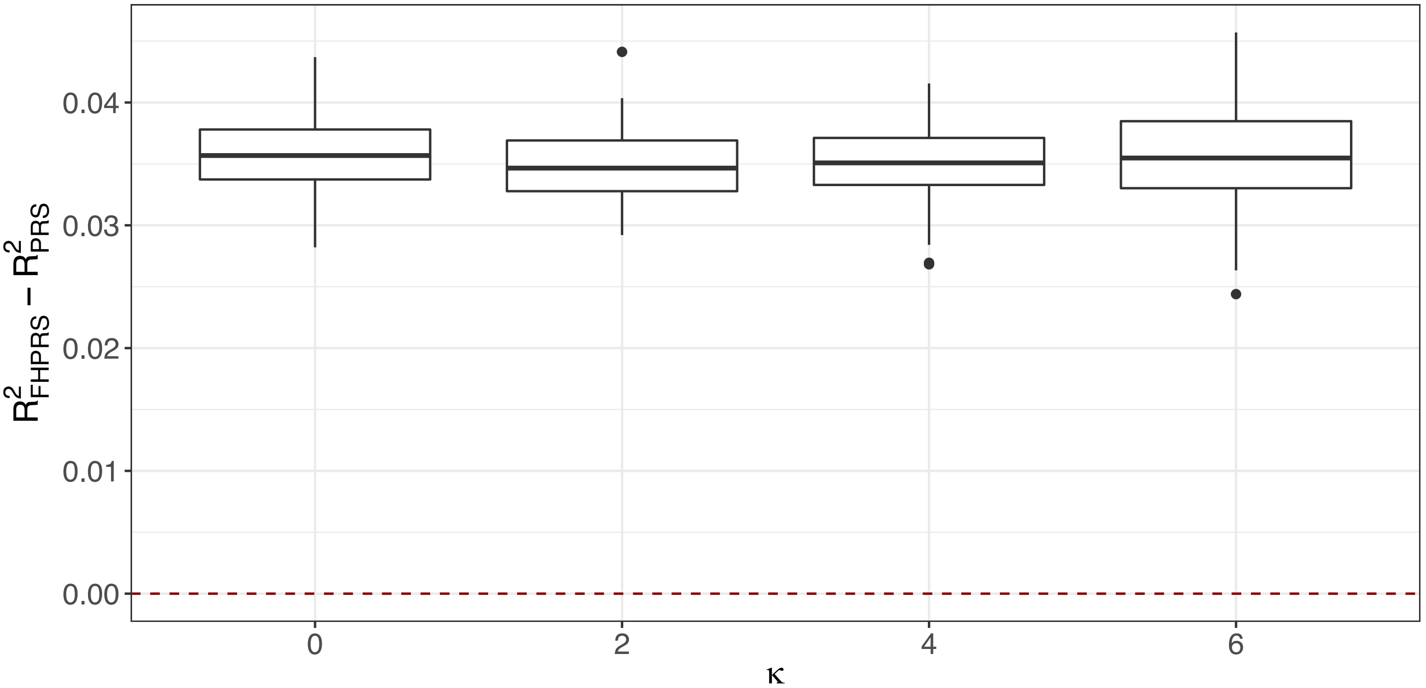


Figure S10. Impact of rare pathogenic variants in the under-modelled genetic component on model performance. The difference in proportion of variance explained for the simulated continuous trait based on the test dataset was calculated with different numbers of rare pathogenic variants (Supplementary Notes). $\kappa = 0$ represents simulation settings with normally distributed under-modelled genetic component. All simulation settings were repeated 100 times with $\alpha = 0.5$ and $\beta = 0.5$ to obtain the distributions of the difference in proportion of variance explained, represented by box plots. FHPRS: family history-assisted polygenic risk score; PRS: polygenic risk score.

Table S1. Cohort characteristics of the Avon Longitudinal Study of Parents and Children.

| | **Women** | **Men** |
|---|---|---|
| Children with mid-parental height available (N = 941) | | |
| Sample size (%) | 541 (57.5) | 400 (42.5) |
| Mean height in cm (SD) | 166.5 (6.0) | 180.1 (6.8) |
| Children with maternal height available[a] (N = 2,246) | | |
| Sample size (%) | 1,352 (60.2) | 894 (39.8) |
| Mean height in cm (SD) | 166.4 (6.0) | 180.1 (6.7) |
| Children with paternal height available[a] (N = 1,092) | | |
| Sample size (%) | 628 (57.5) | 464 (42.5) |
| Mean height in cm (SD) | 166.5 (6.0) | 180.1 (6.7) |

[a] Including 941 children with mid-parental height available

Table S2. Cohort characteristics of the samples from UK Biobank used in this study.

| | **UK Biobank training dataset (10%)** | **UK Biobank test dataset (90%)** |
|---|---|---|
| Mean age (SD) | 56.7 (8.0) | 56.7 (8.0) |
| % Men[a] | 44.8 | 44.9 |
| Breast cancer (N = 230,240) | | |
| Diagnosis (%) | 1,379 (5.9) | 12,476 (6.0) |
| Maternal disease history (%) | 1,840 (7.9) | 16,911 (8.2) |
| Prostate cancer (N = 184,042) | | |
| Diagnosis (%) | 979 (5.3) | 8,806 (5.3) |
| Paternal disease history (%) | 1,317 (7.2) | 12,220 (7.4) |
| Colorectal cancer (N = 393,236) | | |
| Diagnosis (%) | 721 (1.8) | 6,528 (1.8) |
| Maternal disease history (%) | 1,922 (4.9) | 17,339 (4.9) |
| Paternal disease history (%) | 2,126 (5.4) | 19,754 (5.6) |
| Lung cancer (N = 393,236) | | |
| Diagnosis (%) | 377 (1.0) | 3,324 (0.9) |
| Maternal disease history (%) | 1,495 (3.8) | 13,744 (3.9) |
| Paternal disease history (%) | 3,333 (8.5) | 30,755 (8.7) |
| Myocardial infarction (N = 399,318) | | |
| Diagnosis (%) | 1,134 (2.8) | 9,882 (2.7) |
| Maternal disease history (%) | 7,769 (19.5) | 69,004 (19.2) |
| Paternal disease history (%) | 12,226 (30.7) | 110,935 (30.9) |
| Ischemic heart disease (N = 399,318) | | |
| Diagnosis (%) | 4,064 (10.2) | 36,137 (10.1) |
| Maternal disease history (%) | 7,769 (19.5) | 69,004 (19.2) |
| Paternal disease history (%) | 12,226 (30.7) | 110,935 (30.9) |
| Stroke (N = 399,318) | | |
| Diagnosis (%) | 837 (2.1) | 8,042 (2.2) |
| Maternal disease history (%) | 5,472 (13.7) | 49,044 (13.6) |
| Paternal disease history (%) | 5,874 (14.7) | 51,999 (14.5) |
| Type 2 diabetes (N = 399,318) | | |
| Diagnosis (%) | 2,612 (6.6) | 23,435 (6.5) |
| Maternal disease history (%) | 3,459 (8.7) | 31,811 (8.8) |
| Paternal disease history (%) | 3,529 (8.9) | 31,767 (8.8) |
| Alzheimer’s disease (N = 399,318) | | |
| Diagnosis (%) | 180 (0.5) | 1,570 (0.4) |
| Maternal disease history (%) | 3,400 (8.5) | 29,719 (8.3) |
| Paternal disease history (%) | 1,817 (4.5) | 15,991 (4.4) |
| Parkinson’s disease (N = 393,236) | | |
| Diagnosis (%) | 220 (0.6) | 2,050 (0.6) |
| Maternal disease history (%) | 597 (1.5) | 5,523 (1.6) |
| Paternal disease history (%) | 940 (2.4) | 8,325 (2.4) |
| COPD (N = 399,318) | | |
| Diagnosis (%) | 1,476 (3.7) | 13,009 (3.6) |
| Maternal disease history (%) | 2,221 (5.6) | 20,258 (5.6) |
| Paternal disease history (%) | 4,289 (10.7) | 38,634 (10.8) |

[a] Except for breast cancer and prostate cancer studies

Table S3. Information about the polygenic risk scores used for each trait and disease.

| Trait | PGS Catalog[a] ID | Source population for developing score | Method for developing score | Number of predictor variants |
|---|---|---|---|---|
| Height | PGS000758 | European ancestry populations included in GWAS meta-analysis | Hard-thresholding regularized regression | 33,938 |
| Breast cancer | PGS000004 | European ancestry populations included in GWAS meta-analysis | Hard-thresholding stepwise forward regression | 313 |
| Prostate cancer | PGS000030 | European ancestry populations included in GWAS meta-analysis | Genome-wide significant SNPs | 147 |
| Colorectal cancer | PGS000148 | European ancestry populations included in GWAS meta-analysis | Multivariate logistic regression based on genome-wide significant SNPs | 63 |
| Lung cancer | PGS000070 | Populations of European and East Asian ancestries included in GWAS meta-analysis | Genome-wide significant SNPs | 19 |
| Myocardial infarction | PGS000018 | Populations of diverse ancestries included in GWAS meta-analysis | Meta-analysis of polygenic risk scores | 1,745,180 |
| Ischemic heart disease | PGS000018 | Populations of diverse ancestries included in GWAS meta-analysis | Meta-analysis of polygenic risk scores | 1,745,180 |
| Stroke | PGS000039 | Populations of diverse ancestries included in GWAS meta-analysis | Meta-analysis of polygenic risk scores | 3,225,583 |
| Type 2 diabetes | PGS000014 | European ancestry populations included in GWAS meta-analysis | LDPred | 6,917,436 |
| Alzheimer's disease | PGS000026 | European ancestry populations included in GWAS meta-analysis | Hazard model with stepwise selection | 31 |
| Parkinson's disease | PGS000056 | European ancestry populations included in GWAS meta-analysis | Genome-wide significant SNPs | 23 |
| COPD | PGS000210 | European ancestry populations included in GWAS meta-analysis | Genome-wide significant sentinel variants | 279 |

[a] Publicly available at https://www.pgscatalog.org/

Table S4. Model parameter estimates.

| **Continuous trait** | | | | | | | | |
|---|---|---|---|---|---|---|---|---|
| | **Proportion of trait variance captured by polygenic risk score**[a] | **Proportion of variance explained by mid-parental height**[b] | | $\hat{\boldsymbol{\alpha}}$ | $\hat{\boldsymbol{\beta}}$ | | | **Proportion of trait variance not captured by polygenic risk score** |
| Height z-score | 0.367 | 0.449 | | 0.606 | 0.762 | | | 0.581 |
| **Binary disease outcome**[c] | | | | | | | | |
| | **Odds ratio associated with one standard deviation increase in polygenic risk score (95% CI; p-value)** | **Odds ratio associated with maternal disease history (95% CI; p-value)** | **Odds ratio associated with paternal disease history (95% CI; p-value)** | $\hat{\boldsymbol{\alpha}}$ | $\hat{\boldsymbol{\beta}}$ | $\hat{\boldsymbol{\mu}}_{\boldsymbol{0}}$[d] | **Proportion of trait variance captured by polygenic risk score**[e] | **Proportion of trait variance not captured by polygenic risk score**[e] |
| Breast cancer | 1.69 (1.60-1.79; 1.6x10$^{-77}$) | 1.61 (1.35-1.91; 7.4x10$^{-8}$) | | 0.526 | 0.973 | -4.717 | 0.063 | 0.197 |
| Prostate cancer | 1.84 (1.73-1.97; 9.1x10$^{-76}$) | | 2.02 (1.64-2.48; 2.2x10$^{-11}$) | 0.611 | 1.229 | -9.709 | 0.038 | 0.144 |
| Colorectal cancer | 1.22 (1.13-1.31; 1.5x10$^{-7}$) | 1.25 (0.93-1.69; 1.3x10$^{-1}$) | 1.59 (1.22-2.08; 5.6x10$^{-4}$) | 0.197 | 0.833 | -9.231 | 0.004 | 0.074 |
| Lung cancer | 1.14 (1.03-1.26; 1.2x10$^{-2}$) | 1.28 (0.81-2.03; 2.9x10$^{-1}$) | 1.74 (1.31-2.31; 1.5x10$^{-4}$) | 0.130 | 0.884 | -20.775 | 0.001 | 0.036 |
| Myocardial infarction | 1.68 (1.58-1.79; 4.4x10$^{-64}$) | 1.48 (1.29-1.70; 1.4x10$^{-8}$) | 1.49 (1.32-1.68; 2.7x10$^{-10}$) | 0.521 | 0.827 | -6.988 | 0.040 | 0.096 |
| Ischemic heart disease | 1.47 (1.42-1.52; 8.6x10$^{-109}$) | 1.68 (1.55-1.81; 1.1x10$^{-39}$) | 1.52 (1.42-1.63; 6.4x10$^{-32}$) | 0.388 | 1.122 | -7.191 | 0.022 | 0.165 |
| Stroke | 1.24 (1.16-1.33; 5.4x10$^{-10}$) | 0.97 (0.80-1.18; 7.9x10$^{-1}$) | 1.14 (0.95-1.37; 1.6x10$^{-1}$) | 0.219 | 0.242 | -8.000 | 0.006 | 0.008 |
| Type 2 diabetes | 1.65 (1.58-1.72; 1.3x10$^{-123}$) | 2.64 (2.37-2.95; 4.1x10$^{-67}$) | 2.29 (2.04-2.57; 3.0x10$^{-44}$) | 0.401 | 2.000 | -6.462 | 0.040 | 0.439 |
| Alzheimer's disease | 1.19 (1.02-1.38; 2.3x10$^{-2}$) | 1.98 (1.36-2.88; 3.7x10$^{-4}$) | 1.16 (0.61-2.20; 6.6x10$^{-1}$) | 0.172 | 0.894 | -26.597 | 0.001 | 0.028 |
| Parkinson's disease | 1.36 (1.20-1.55; 2.6x10$^{-6}$) | 1.59 (0.70-3.62; 2.7x10$^{-1}$) | 2.06 (1.11-3.80; 2.1x10$^{-2}$) | 0.309 | 1.044 | -24.250 | 0.004 | 0.042 |
| COPD | 1.10 (1.04-1.16; 4.8x10$^{-4}$) | 2.48 (2.10-2.92; 2.5x10$^{-27}$) | 1.73 (1.51-1.98; 5.8x10$^{-15}$) | 0.096 | 1.203 | -18.964 | 0.001 | 0.071 |

[a] Based on the UK Biobank

[b] Based on the Erasmus Rucphen Family Study

[c] Based on the UK Biobank training dataset

[d] Effects of age and sex were modeled as covariate effects and were not captured in $\hat{\mu}_0$

[e] Total trait variance includes covariate effects

Table S5. Summary of the area under the precision-recall curve based on the UK Biobank test dataset.

| **Disease** | **AUPRC**[a] | | |
|---|---|---|---|
| | Polygenic risk score | Parental history | Joint predictor |
| Breast cancer | 0.1025 | 0.0769 | 0.1058 |
| Prostate cancer | 0.1485 | 0.1022 | 0.1496 |
| Colorectal cancer | 0.0346 | 0.0332 | 0.0354 |
| Lung cancer | 0.0201 | 0.0205 | 0.0210 |
| Myocardial infarction | 0.0800 | 0.0671 | 0.0834 |
| Ischemic heart disease | 0.2464 | 0.2341 | 0.2562 |
| Stroke | 0.0456 | 0.0434 | 0.0459 |
| Type 2 diabetes | 0.1487 | 0.1485 | 0.1634 |
| Alzheimer's disease | 0.0130 | 0.0135 | 0.0142 |
| Parkinson's disease | 0.0158 | 0.0148 | 0.0162 |
| COPD | 0.0945 | 0.1044 | 0.1057 |

[a] Including effects of age, sex, genotyping array, recruitment centre, and the first 10 genetic principal components
